# Multi-modal single-cell sequencing identifies cellular immunophenotypes associated with juvenile dermatomyositis disease activity

**DOI:** 10.1101/2021.09.18.21263581

**Authors:** Jessica Neely, George Hartoularos, Daniel Bunis, Yang Sun, David Lee, Susan Kim, Chun Jimmie Ye, Marina Sirota

**Affiliations:** Division of Pediatric Rheumatology, Department of Pediatrics, University of California San Francisco School of Medicine; Graduate Program in Biological and Medical Informatics, University of California, San Francisco; Institute of Human Genetics, University of California, San Francisco; Division of Rheumatology, Department of Medicine, University of California, San Francisco; UCSF CoLabs, University of California, San Francisco; ImmunoX Initiative, University of California, San Francisco; Department of Epidemiology and Biostatistics, University of California, San Francisco; Bakar Computational Health Sciences Institute, University of California, San Francisco; Parker Institute for Cancer Immunotherapy; Chan Zuckerberg Biohub; Department of Pediatrics, University of California, San Francisco

## Abstract

Juvenile dermatomyositis (JDM) is a rare autoimmune condition with insufficient biomarkers and treatments, in part, due to incomplete knowledge of the cell types mediating disease. We investigated immunophenotypes and cell-specific genes associated with disease activity using multiplexed RNA and protein single-cell sequencing applied to PBMCs from 4 treatment-naïve JDM (TN-JDM) subjects at baseline, 2, 4, and 6 months and 4 subjects with inactive disease. Analysis of 55,564 cells revealed separate clustering of TN-JDM cells within monocyte, NK, CD8+ effector T and naïve B populations. The proportion of CD16+ monocytes was reduced in TN-JDM, and naïve B cells were expanded. Cell-type differential gene expression analysis and hierarchical clustering identified a pan-cell-type IFN gene signature over-expressed in TN-JDM in all cell types and correlated with disease activity. TN-JDM monocytes displayed an inflammatory state: CD16+ monocytes expressed the highest IFN gene score and differential protein expression of adhesion molecules, CD49d and CD56, compared to CD14+ inflammatory monocytes. A transitional B cell population expressing higher CD24 and CD5 proteins and an IFN-hi naïve B population were associated with TN-JDM and exhibited less CD39, an immunoregulatory protein. This data provides new insights into JDM immune dysregulation at cellular resolution and novel resource for myositis investigators.

## INTRODUCTION

Juvenile dermatomyositis (JDM) is a complex immune-mediated disease characterized by inflammatory myopathy of proximal musculature and pathognomonic skin rashes. Immune mechanisms are not fully understood, however, investigations to date implicate a combination of genetic variants and environmental influences (1). Though mortality rates have greatly improved, more than 60% of children with JDM have long-term damage and functional impairment due to poorly controlled disease or corticosteroid toxicity (2). This is, in part, due to the paucity of reliable, objective biomarkers to monitor disease activity, predict flares, or guide treatment decisions and an incomplete understanding of the cellular immunophenotypes contributing to disease. To improve the outcomes of children with JDM, a personalized approach to disease management, is urgently needed.

A key step in developing a personalized treatment strategy in JDM is a better understanding of the interferon (IFN) response, including the specific cell types involved in IFN signaling in JDM and the dynamic nature of the response during the course of disease. Numerous studies investigating gene expression in adult dermatomyositis (DM) and JDM have shown that the transcriptional signature is dominated by type I IFN stimulated genes (ISGs) (3–6). Some, but not all studies, have shown a correlation with ISG expression and disease activity (3,7,8). This discrepancy is likely due to the limitations of prior expression profiling technology, which measure transcriptional changes across aggregated cell types, as well as nuances of IFN signaling, which have been shown to be both cell-type-specific and disease-specific (9). Single-cell sequencing, using a multi-modal approach measuring gene and protein expression, represents a unique opportunity to characterize immune cell types and cell-specific IFN responses in JDM, in an unbiased fashion.

In addition to characterizing the cell-type specific IFN response in JDM, a better understanding of the cell types that contribute to disease is imperative. Immune mechanisms in JDM are complex, involving both innate and adaptive arms of the immune system, as well as many cell types (1). A role for B cells has been supported by the presence of myositis-specific antibodies that correlate with distinct clinical phenotypes (10) and by a positive clinical response to rituximab, a B-cell depleting agent (11). Likewise, two independent studies have also demonstrated an expansion of naïve B cells in JDM (12, 13). However, the pathogenicity of these antibodies or how naïve B cells contribute to disease is still unclear. Several studies have demonstrated infiltration of T cells in muscle and skin biopsies in DM/JDM using immunohistochemistry (14–17), and flow cytometry studies demonstrated a skewing of peripheral blood CXCR5+Th subsets toward Th2 and Th17 phenotypes with increasing disease activity (18). Additionally, peripheral blood NK cells are decreased in number and dysfunctional, suggesting a role for innate immune cells as well (13).

In this study, we sought to build upon this knowledge by comprehensively interrogating the peripheral blood compartment in JDM, including simultaneous measurements of RNA and cell-surface proteins. Our goal was to provide an unbiased characterization of peripheral blood immune cells as well as insight into the dynamic nature of immune cell composition and cell-specific transcriptomic and proteomic signatures during the course of treatment. This work lays a foundation for future cell-specific biomarker and drug target development to help myositis researchers develop and implement a personalized treatment strategy to improve the outcomes of children with JDM.

## RESULTS

### JDM is associated with alterations in immune cell composition

A graphical depiction of our study design, multiplexing strategy, and analysis pipeline is depicted in Figure 1. Four subjects with JDM with treatment-naïve disease (TN-JDM) and 4 subjects with established, clinically inactive disease (I-JDM) were included. Serial samples from TN-JDM subjects were included at baseline and approximately, 2, 4, and 6 months into treatment. The median age of JDM diagnosis was 9.5 years in the TN-JDM group and 7 years in the I-JDM group, and the median age at study enrollment in both groups was 9.5 years (Table 1). Two of four patients were female in the TN-JDM group and 3 out of 4 were female in the I-JDM group. In both groups, 2/4 patients were positive for TIF1-γ. All patients exhibited improvement in disease activity measures during the first 6 months of disease (Table 1 and Supplementary Figure 1).

**Figure 1:**
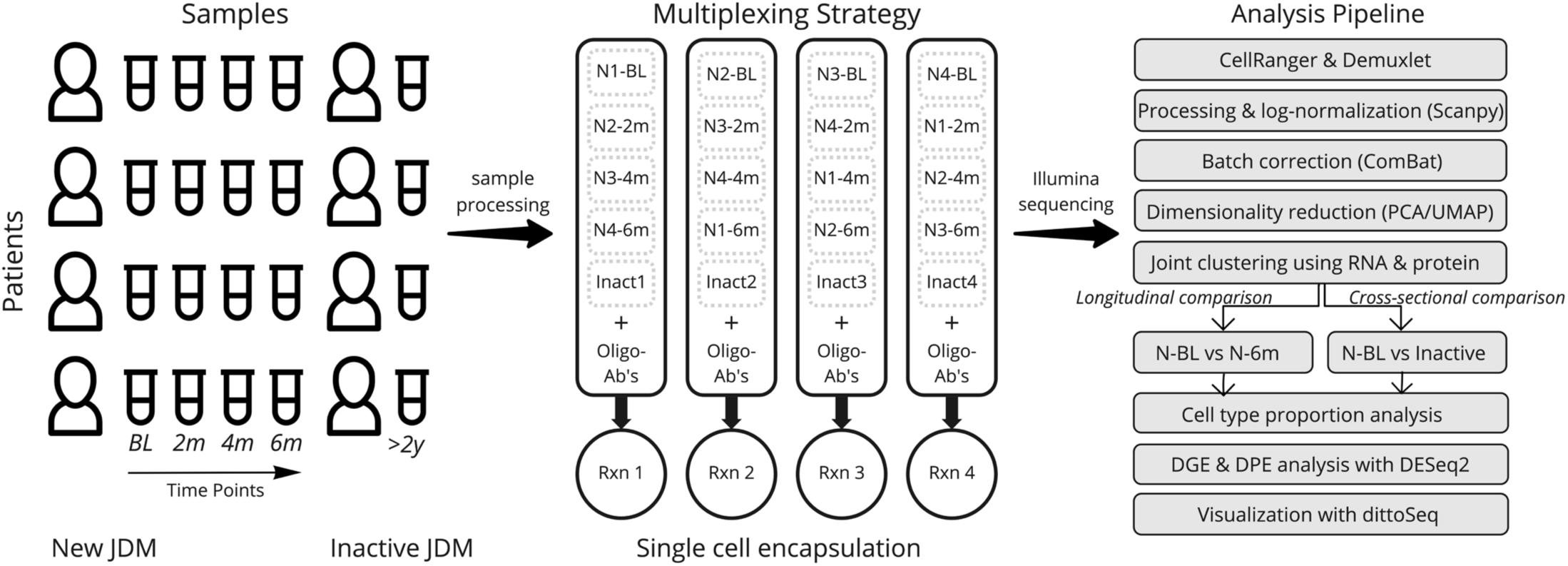
Experimental Overview. An overview of the experimental design, multiplexing strategy, and analysis pipeline. Timepoints refer to longitudinal samples obtained from the same patients at BL=baseline, 2m=2 months, 4m=4 months, 6m=6 months; N refers to “New-JDM”. Rxn= reaction, DGE=differential gene expression, DPE=differential protein expression.

**Table 1:** Patient Characteristics.

|  | I-JDM | TN-JDM |  |  |  |
| --- | --- | --- | --- | --- | --- |
|  |  | Baseline | 2 months | 4 months | 6 months |
| Median age (yrs) at diagnosis | 7 | 9.5 | - | - | - |
| Median age (yrs) at study visit | 9.5 | 9.5 | - | - | - |
| Female sex (%) | 75% | 50% | - | - | - |
| TIF1-y (%) | 50% | 50% | - | - | - |
| NXP-2 (%) | 25% | 25% | - | - | - |
| MDA-5 (%) | - | 25% | - | - | - |
| MSA-neg (%) | 25% | - | - | - | - |
| Median disease duration (mos) | 38.2 | 2.7 | - | - | - |
| Physician VAS Global (median [IQR]) | 0.4<br>[0.2, 0.5] | 5.5<br>[4.6, 6.0] | 2.3<br>[1.6, 2.9] | 1.3<br>[0.8, 1.6] | 1.0<br>[0.4, 1.5] |
| Physician VAS Cutaneous (median [IQR]) | 0.3<br>[0.0, 0.5] | 4.5<br>[3.1, 5.9] | 2.3<br>[1.6, 2.6] | 1.8<br>[1.3, 2.0] | 1.3<br>[0.8, 1.6] |
| Physician VAS Muscle (median [IQR]) | 0.2<br>[0.0, 0.5] | 4.0<br>[3.3, 4.0] | 0.8<br>[0.4, 1.0] | 0.0<br>[0.0, 0.1] | 0.0<br>[0.0, 0.1] |
| MMT-8 (median [IQR]) | 79.0<br>[78.0, 80.0] | 70.0<br>[69.5, 73.0] | 80.0<br>[78.5, 80.0] | 80.0<br>[79.0, 80.0] | 80.0<br>[79.5, 80.0] |
| CHAQ Disability Index (median [IQR]) | 0.00<br>[0.00, 0.09] | 0.62<br>[0.62, 1.25] | 0.31<br>[0.22, 0.69] | 0.31<br>[0.09, 0.56] | 0.06<br>[0.00, 0.19] |
| CDASI Activity Score (median [IQR]) | 1.0<br>[0.0, 2.3] | 21.5<br>[13.0, 29.0] | 5.5<br>[3.3, 7.8] | 4.5<br>[3.3, 5.0] | 3.0<br>[1.8, 4.3] |

Abbreviations: I-JDM = inactive juvenile dermatomyositis, TN-JDM = treatment-naïve juvenile dermatomyositis, MSA-neg = myositis-specific antibody negative, VAS = visual analog scale, IQR = interquartile range, MMT-8 manual muscle testing 8, CHAQ = childhood health assessment questionnaire, CDASI cutaneous dermatomyositis disease area and severity index

After filtering and doublet removal, we analyzed 55,564 cells comprising all the major immune cell populations, which were annotated using canonical cell markers (Figure 2a). Cells from TN-JDM subjects clustered together and occupied distinct regions of UMAP embeddings in CD14+ and CD16+ monocyte, naïve B cell, naïve and effector CD4+ and CD8+ T cell and CD56dim NK cell populations (Fig. 2b). Visualization of UMAP embeddings by visit number demonstrated a shift in embeddings over time from tightly clustered regions in TN-DM to a broader embedding across the cluster at subsequent time points suggesting diversification of cell states as disease activity declines with treatment (Fig. 2b). Cells from the 4 month and 6 months visits occupied similar regions as cells from I-JDM in most immune cell populations.

**Figure 2:**
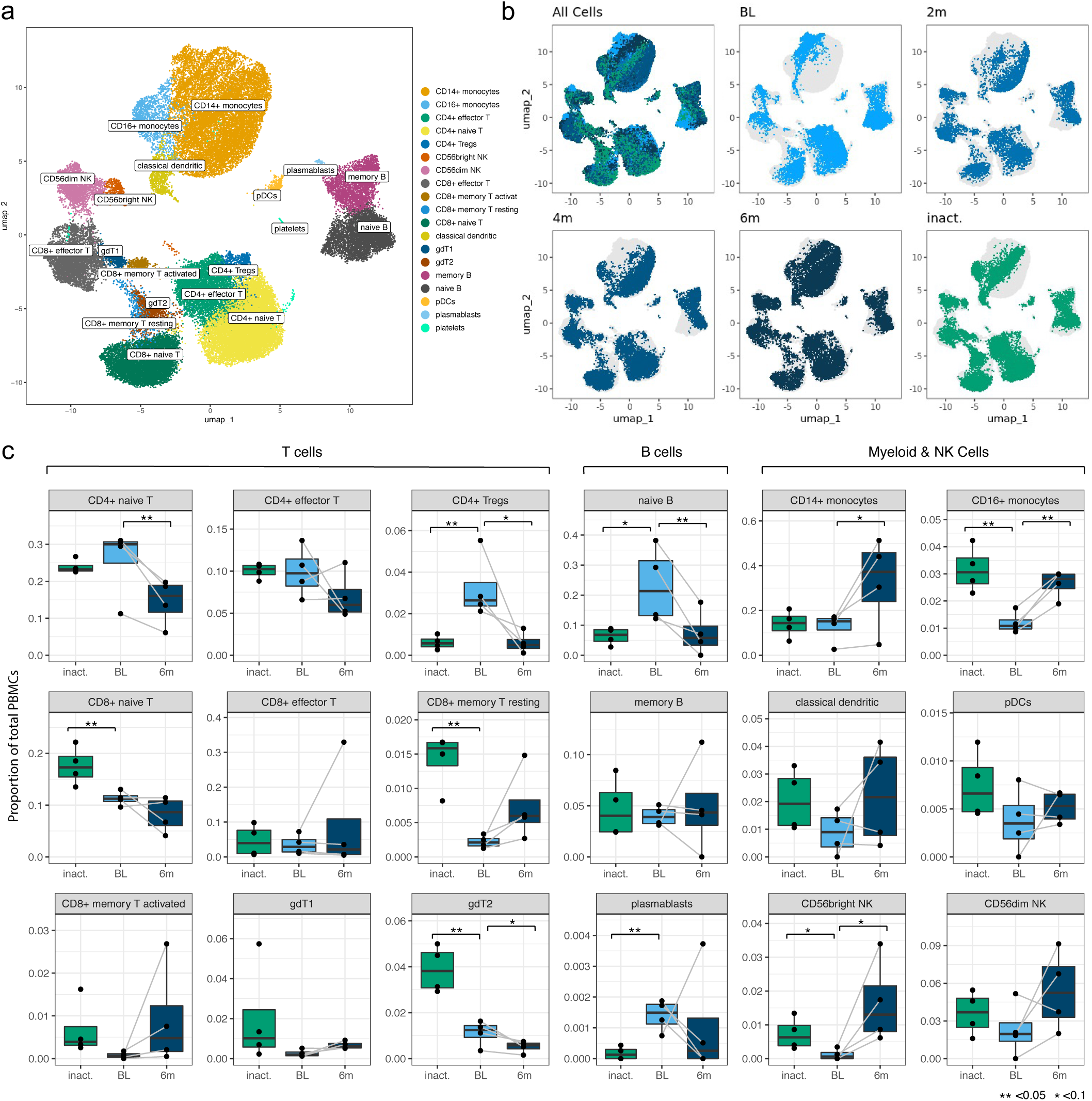
TN-JDM cells display alterations in cell composition compared to treated JDM. **a.** A UMAP plot showing 19 manually annotated cell types from 24 immune cell populations identified by Leiden clustering. Manual annotations were based on canonical markers shown in Supplementary Figure 2 **b.** UMAP embeddings showing all cells colored by timepoint (for newly-diagnosed patients) and inactive disease state showing distinct regions of embedding for cells from BL TN (treatment-naïve) patients in most cell populations. **c.** Boxplots for 18 cell types (platelets not included) showing differences in cell type proportion between BL TN samples and inactive samples and between BL, TN samples and 6m samples. Comparison of proportions between visit 1 and visit 4 was a paired analysis and grey lines connect the observations from the same individuals. P values were calculated using a T test (**<0.05, *<0.1).

Cell type proportions were altered in TN-JDM compared to treated JDM (Fig. 2c). In a paired analysis of TN-JDM comparing baseline samples to 6 month samples, there was a significant expansion of naïve B cells (p=0.03) and CD4+ naïve T cells (p=0.02) and a reduction of CD16+ monocytes (p=0.01). In one patient, subsequent reduction of naïve B cells at the 6 month visit was medication-induced due to treatment with a B-cell depleting agent, rituximab. In a cross-sectional analysis comparing TN-JDM to I-JDM, there was also a reduction of CD16+ monocytes (p=0.01) and trend toward expansion of naïve B cells (p=0.07). There were also significant differences in several T cell populations with expansion of CD4+ Tregs (p=0.04) and a reduction of CD8+ naïve T cells (p=0.04), and gdT2 populations (p=0.01). The increase in the CD8+ memory T resting (p=0.01) population at visit 4 and to higher levels in inactive JDM suggests a memory T cell response that develops over time in JDM. In both analyses, there was a trend toward reduction of CD56bright NK cells (p=0.07 in paired, and p=0.08 cross-sectionally). Expansion of naïve B cells, and alterations in regulatory cell types, including CD16+ monocytes and CD4+Tregs, may be associated with active JDM.

### Global and cell type-specific transcriptomic signatures in JDM are associated with disease activity

To develop a global understanding of the JDM transcriptional signature and how it changes with treatment, we first performed differential gene expression analysis between TN-JDM baseline and 6 months visits and between TN-JDM and I-JDM groups (Supplementary Table 1). Monocytes, classical dendritic cells (cDCs), and cytotoxic cell types, CD8+ effector T and CD56dim NK cells, displayed the greatest number of differentially expressed genes (DEGs) and many genes were differentially expressed in multiple cell types (Supplementary Figures 6 and 7). Unsupervised clustering using cell-specific DEGs clustered patients according to disease activity levels, as expected (Supplementary Figs. 8-13). We then calculated the pseudobulk expression profiles per disease group (baseline, 2 mos, 4 mos, 6 mos, and inactive) for 368 genes that were differentially expressed in at least one cell type in both analyses (Fig 3a). Hierarchical clustering of this gene set revealed 10 gene modules, which were visualized using a heatmap with columns ordered by descending level of disease activity (baseline, 2 mos, 4 mos, 6 mos, and inactive) and cell type. Gene ontology enrichment analysis of each of the ten gene modules identified associations with protein translation, cytokine regulation, type I and II interferon signaling, NFκB signaling, antigen presentation via the class I pathway, and the unfolded protein response (Fig. 3b & Supplementary Fig. 14). Module 9 did not have any significant enrichment terms, but these genes were highly expressed in plasmacytoid dendritic cells (pDCs). Three modules were enriched in IFN signaling, including type I and type II IFN responses: modules 3, 5, and 8 (Fig. 3b). Module 8 genes were highly expressed in the TN-JDM group in nearly every cell type, and expression sharply declined with treatment. This “Pan-cell IFN” signature was most prominently expressed in CD16+ and CD14+ monocytes, CD56dim NK cells and CD8+ effector T cells in TN-JDM subjects (Supplementary Fig. 15). In contrast, module 3 genes were more prominently expressed in myeloid cell types and module 5 genes were more prominently expressed in T and NK cell types, and gene expression within these cell types displayed a more static expression pattern over time for the respective IFN gene modules. CD16+ monocytes displayed the highest gene scores in TN-JDM patients for both the myeloid IFN module and the “Pan-cell IFN” module (Supplementary Fig. 15).

**Figure 3:**
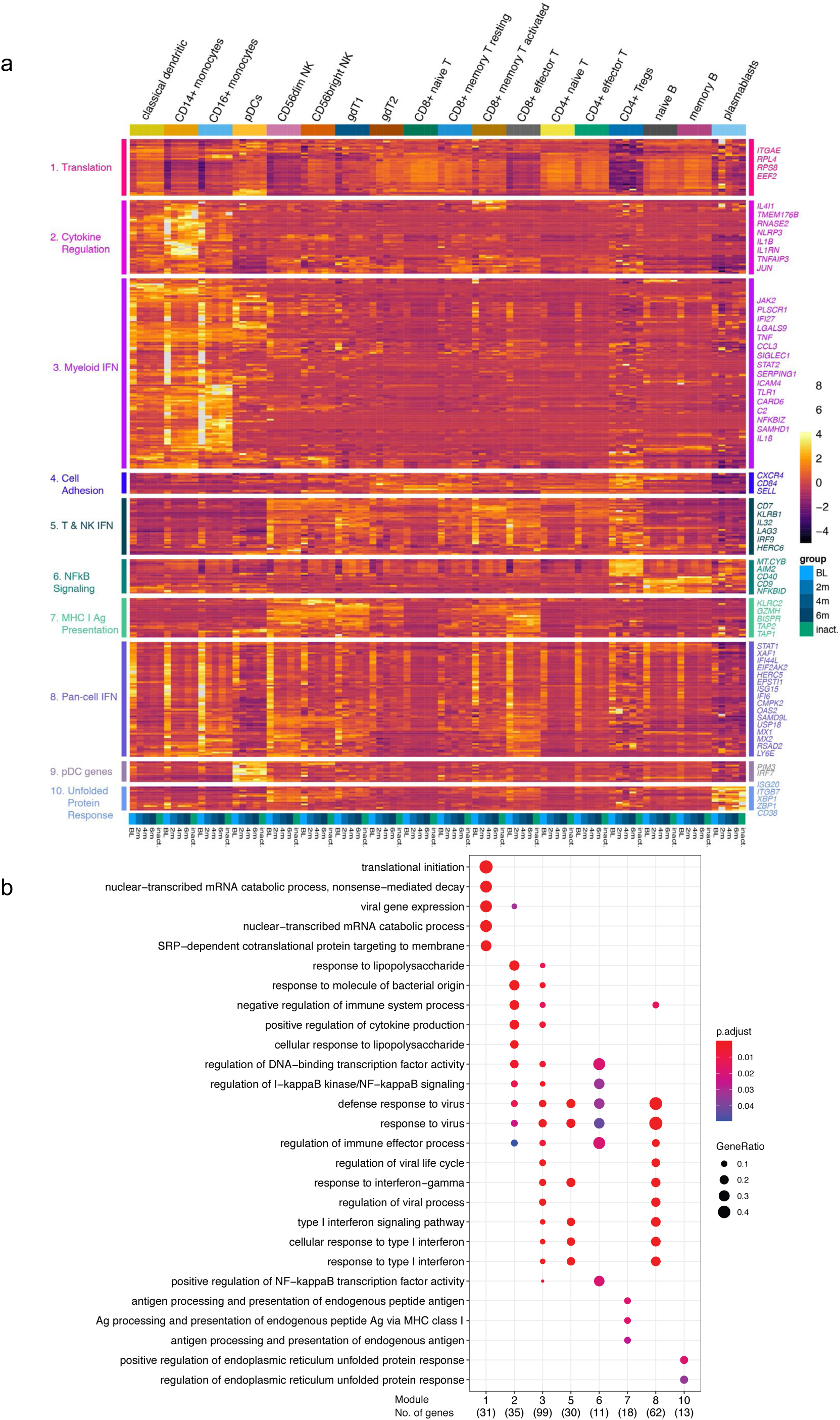
The IFN response in JDM is both broadly expressed and cell-specific. **a**. A heatmap displaying the mean expression of 368 genes differentially expressed in at least one cell type for each group per cell type ordered by increasing time from diagnosis (BL, 2m, 4m, 6m, inactive). Rows are clustered using unsupervised hierarchical clustering with k=10, and modules are annotated by enrichment terms on the left. A selection of genes from each module is annotated on the right. Color represents the normalized mean expression. **b**. A dotplot displaying the association between each gene module set and enrichment of GO Biologic Processes terms. Each column represents a module from panel a. Enrichment analysis was performed using over-representation analysis with the color representing the adjusted p-value using B&H and the size of the dot representing the ratio of the number of genes relative to the number of genes in each term.

We next wanted to directly determine the association between these gene sets and disease activity measures to identify biologically relevant sets of genes and putative disease activity biomarkers. We calculated a gene score for each module by averaging the expression of the gene set for each sample. The Pearson correlations between the gene score per sample and physician global visual analog score (VAS) score identified a significant correlation in bulk cells for modules 2, 8 and 10 (Fig 4a). Evaluation of the correlations between cell-specific gene scores and disease activity identified stronger correlations in CD56dim NK and CD8 effector T cells for all three modules (Fig 4a and 4b). These cell-specific gene scores were negatively associated with disease activity for module 2 genes and positively correlated with module 8 genes (Fig 4b), which may be suggestive of opposing gene programs that become more regulated with treatment. The magnitude of change in gene scores was greatest within the module 8, “Pan-cell IFN” gene set. Module 10 gene scores displayed very strong correlations in memory and naïve B cells, though the overall magnitude of the change was less. This gene set was also highly expressed in plasmablasts (Fig. 3a). These results help to identify ISG’s expressed more dynamically in JDM from those that display a more static pattern of gene expression in certain cell types and suggests that the pan-cell IFN gene signature might serve as a disease biomarker in JDM in cytotoxic cell types and in bulk cells.

**Figure 4:**
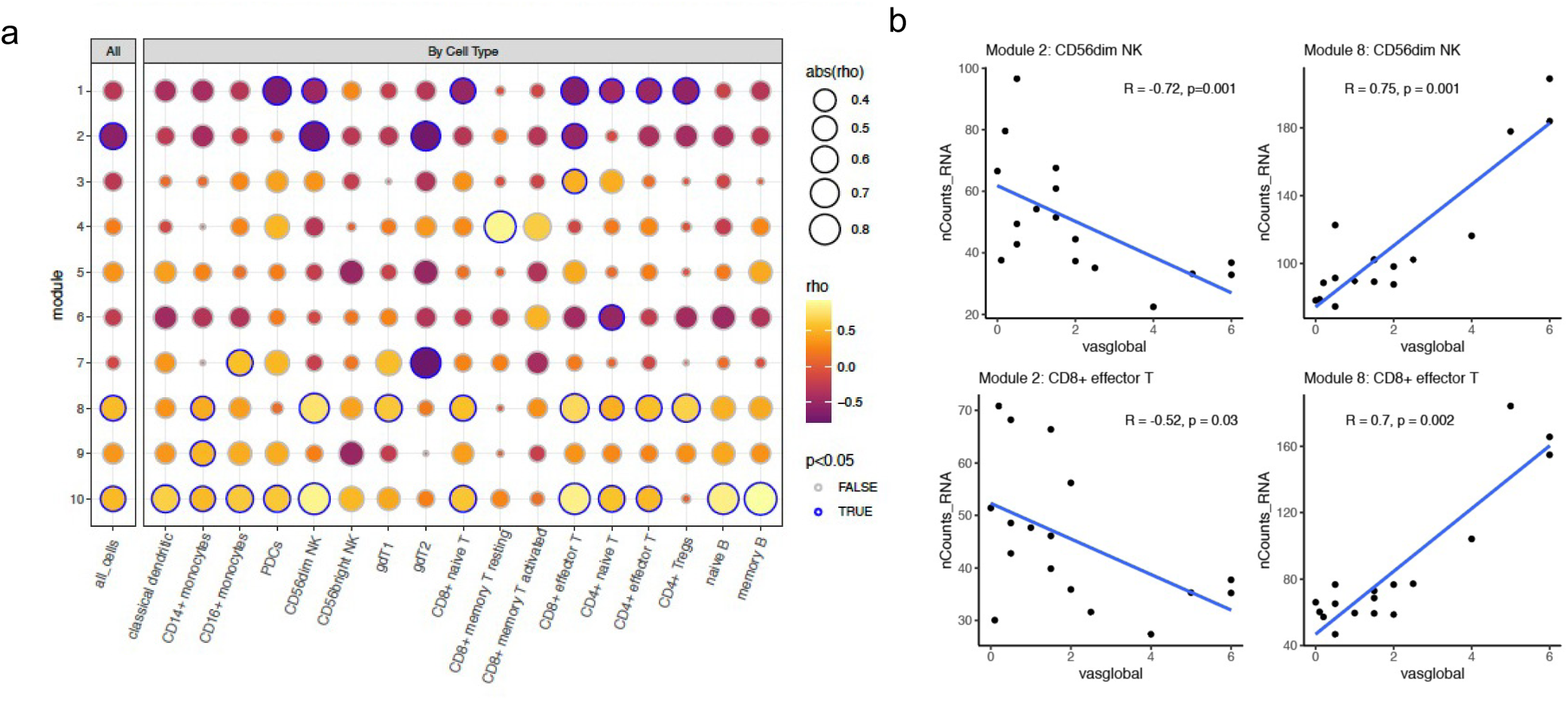
Cytotoxic cell transcriptomic signatures correlate with disease activity. **a**. A dotplot displaying the strength of correlation between each module gene score, calculated as the average expression of all genes within a module, and the physician global VAS score for all cells together (left column) and each cell type individually. The correlations was calculated using a Pearson correlation. The size of each dot represents the strength of the association, the color represents the direction of the association, and the outline of the circle indicates significance defined as P-value <0.05. **b**. The Pearson correlation between the module gene score and physician global VAS plotted for each sample in CD8+ effector T and CD56dim NK cells for modules 2 and 8. The blue line represents a linear model fit for visualization purposes

### Monocytes in TN-JDM display inflammatory and antigen-presenting properties

We next turned our attention to the monocyte populations because of the strong IFN signature, sizeable number of DEGs, and compositional changes observed between disease groups. Differential gene expression results were concordant comparing TN-JDM baseline samples to 6 months samples and TN-JDM to I-JDM in both cell types. TN-JDM subjects displayed up-regulation of IFN-stimulated genes and inflammatory genes, including *NFKBIZ, NFKBIA, IL1B, TNF, CCL3*, and *CCL4* in both monocyte populations (Supplementary Table 1). Despite a reduction in CD16+ monocytes in TN-JDM, this population had the most differentially expressed genes both comparing TN-JDM baseline to 6 mos (n=229) and TN-JDM baseline to I-JDM (n=359) (Supplementary Figures 6 and 7). In addition to ISG’s and inflammatory cytokines, TN-JDM subjects also displayed up-regulation of genes related to antigen presentation (*CD40, HLA.DRB5, HLA.DRB1*, and *TAPBL*), TLR signaling (*TLR1* and *TLR2*), inflammasome signaling (*NLRP3* and *AIM2*), IL-6 signaling (*IL6ST*) and complement genes (*C2, C1QA*), indicating a population with inflammatory and antigen-presenting capabilities. Within the CD14+ monocyte population, TN-JDM subjects also displayed less expression of *CD163* and *IL-18* compared to treated JDM suggesting reduced phagocytic function of the CD14+ monocyte population in active disease. In both monocyte populations, we observed co-expression of genes related to IL-1 signaling and IFN signaling (Fig 5a) that corresponded to the region of embedding of TN-JDM cells. Differential protein analysis within CD14+ monocytes revealed up-regulation of HLA.A-B-C, a marker for MHC class I, in TN-JDM and down-regulation of CD56 and CD11b (Supplementary Table 2). In CD16+ monocytes, there was up-regulation of HLA.A-B-C and CD86, a costimulatory molecule, supporting the antigen-presenting capabilities of these cells at the protein level (Supplementary Table 2). Notably, we observed down-regulation of the surface CD16 in TN-JDM cells.

**Figure 5:**
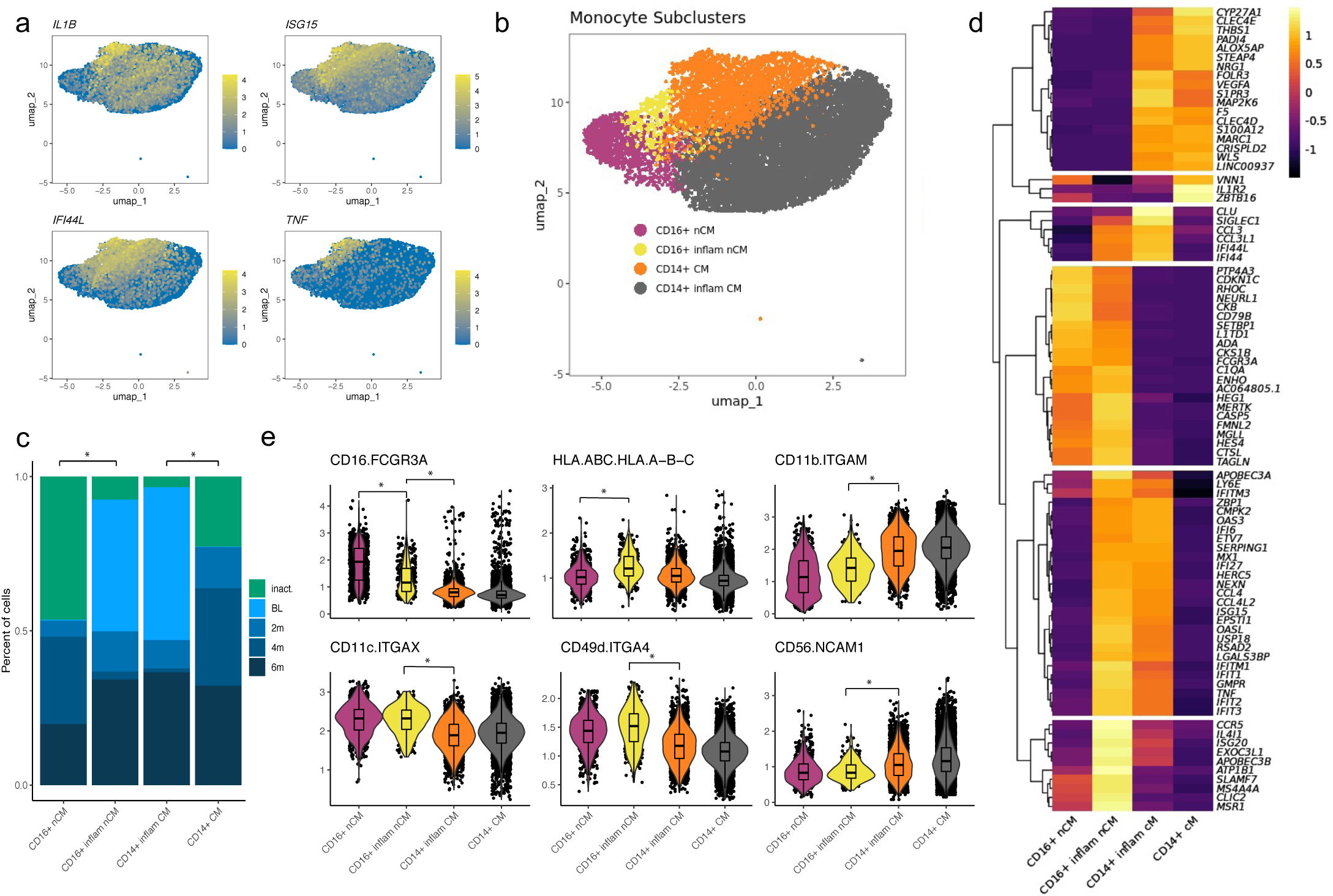
Monocytes in TN-JDM display inflammatory and antigen-presenting properties. **a.** UMAP plots of the monocyte population colored by select features where color represents the log-normalized expression. **b.** UMAP of monocyte populations colored by monocyte cell states following a second round of clustering of CD16+ nCM. **c.** Disease group composition of monocyte subclusters. **d.** A heatmap displaying differentially expressed transcripts between monocyte populations using a log-fold change >3 and *VEGFA* and adjusted p-value <0.05. All genes differentially expressed meeting significance thresholds are displayed in Supplementary Table 2. **e.** Violin plots displaying expression values of differentially expressed proteins between monocyte subsets using a log-fold change >0.5 and adjusted p-value <0.05.

Because of the high number of DEGs and because TN-JDM cells occupied a distinct region within monocytes in the UMAP, we next evaluated the role of monocyte subsets. A second round of clustering was applied to the CD16+ monocyte population and identified a cluster consisting of 98% TN-JDM CD16+ cells (Fig 5b). The composition of this subcluster was significantly different with a high proportion of cells from TN-JDM (43%) as well as cells from 6 month visit, 88% of which were from subject N-1 (Fig 5c). Likewise, 99% of TN-JDM cells in the CD14+ monocyte population resided within original Leiden clusters “10” and “18”, again with 99% of cells from 6 month samples coming from one subject, N-1 (Fig 5c). Both of these subclusters expressed high levels of inflammatory genes like *CCL3, CCL4, TNF* and ISG’s but also retained features of the original CD16+ and CD14+ monocytes, supporting inflammatory cell states of CD16+ and CD14+ monocytes in TN-JDM (Fig 5d). At the gene level CD16+ inflammatory monocytes displayed higher expression of *CCR5* (not recapitulated at the protein level), *IL4IL*, and *ATP1B* than other monocyte subclusters (Fig 5d) and differential expression of many HLA class II genes and CD14+ inflammatory monocytes displayed more S100 protein genes, including *S100A8, S100A9*, and *S100A12* (Supplementary Table 2). At the protein level, CD16+ inflammatory monocytes displayed less CD16+ expression and distinguished from CD14+ monocytes by increased expression of, CD49d, CD16, CD11c and reduced CD11b and CD56 (Fig 5e). While this population displayed less CD16+, *CD14* transcripts were not significantly different between the two CD16+ clusters, and additional canonical markers of intermediate monocytes were not present supporting the classification of this cell type as a CD16+ monocyte population exhibiting an altered inflammatory cell state.

Both inflammatory monocyte populations were embedded within the UMAP further from the classical dendritic cell (cDC) population. This along with the transcriptomic and proteomic features exhibited by these monocyte populations is suggestive of early macrophage differentiation. Interestingly, both CD16+ and CD14+ inflammatory monocytes display features resembling pro-inflammatory M1 macrophages; CD14+ monocytes also display some features resembling anti-inflammatory M2 macrophages, like *VEGFA* and *IL1R2* (decoy receptor). These results support a role for the peripheral blood monocyte compartment in JDM, potentially as precursors to monocyte-derived macrophages in target tissues.

### A transitional B cell population is expanded in JDM

Two populations of B cells (CD19+, CD20+) were identified: naïve (IgD+) and memory B cells (CD27+), as well as a small cluster of plasmablasts (CD27+, CD38+). Naïve B cells were consistently expanded in TN-JDM both longitudinally comparing the baseline visit to 6 month visit and compared to I-JDM. Differential gene expression analysis revealed increased ISG expression in TN-JDM both longitudinally and compared to I-JDM (Supplementary Table 1). Differential protein expression identified down-regulation of CD39 in TN-JDM in both naïve B cells and memory B cells (Supplementary Table 1).

Within the naïve-B cell population, cells from TN-JDM subjects occupied a distinct region (Fig 2b). To better characterize the cell types in this region, a second round of clustering was applied to the naïve B cell population, which identified three subclusters (Fig 6a). These subclusters (SCs) displayed altered composition compared to the naïve B cell cluster with a high proportion of cells from TN-JDM subjects: 64% of SC1 and 87% of SC2 were from TN-JDM subjects (Fig 6b). Differential protein expression analysis comparing each cluster identified higher CD24 and CD5 expression and down-regulation of CD39 in SC1 (Fig 6c). SC2 was differentiated from naïve B cells only by down-regulation of CD39 (Fig 6c). SC1 also expressed surface IgD but not CD27 supporting the classification of this cell type as a transitional B cell population (19, 20) (Fig 6d).

**Figure 6:**
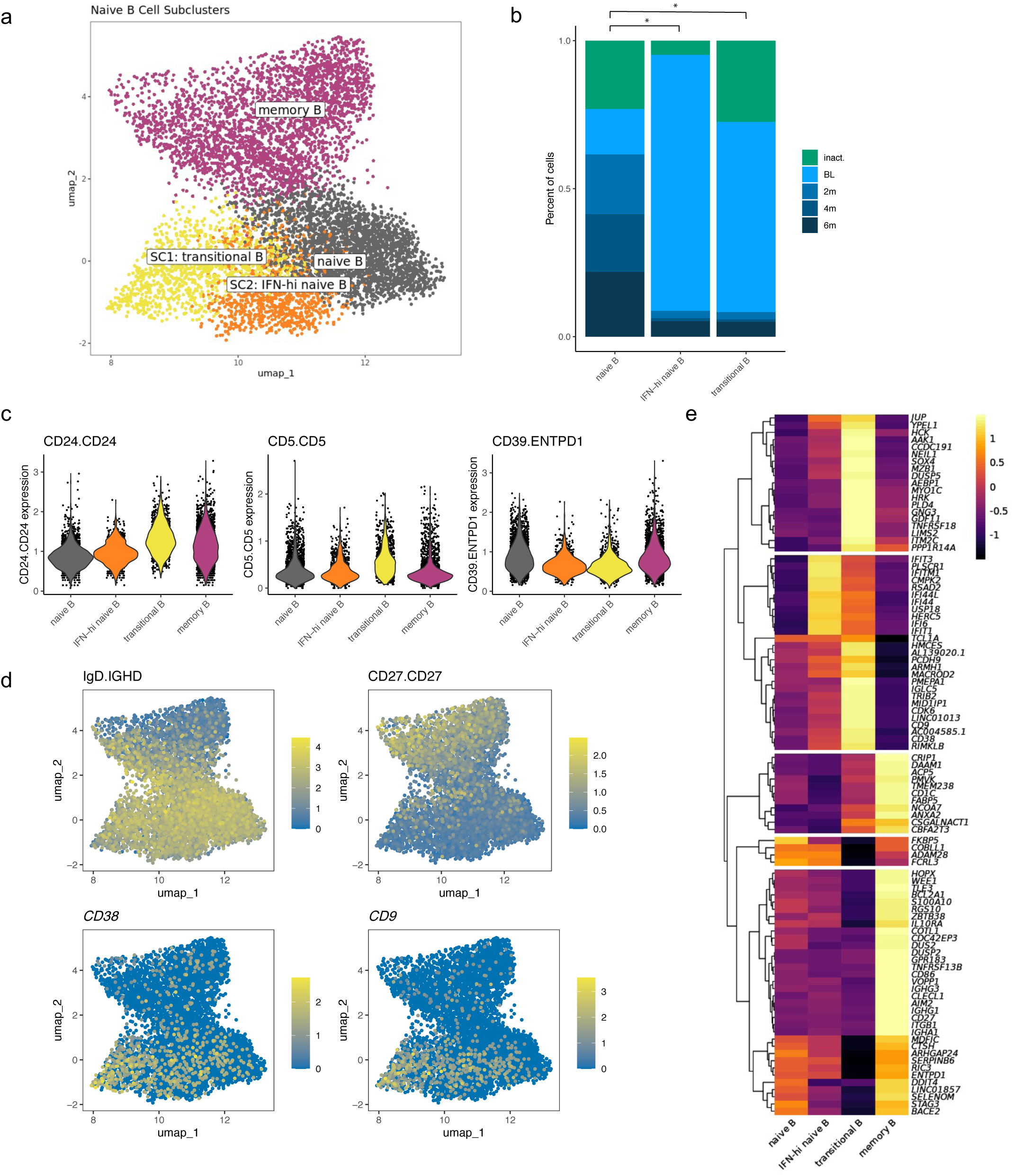
A Transitional B cell population is expanded in JDM. **a**. A UMAP of B cell populations after a second round of naïve B cell clustering colored by naïve B cell subcluster. **b**. Disease group composition of naïve B cell subclusters. Significance is determined using a chi-squared test and p-value <0.05. **c**. Violin plots displaying expression values of differentially expressed proteins between naïve B cell subsets using a log-fold change >0.5 and adjusted p-value <0.05. **d**. UMAPs of B cell populations displaying log-normalized expression of phenotypic markers IgD (protein), CD27 (protein), CD38 (RNA) and CD9 (RNA). **e**. A heatmap displaying differentially expressed transcripts comparing transitional B cells to all other B cells and IFN-high naïve B cells to all other B cells. Genes displayed are those with a log-fold change>1.3, in addition to *IL10RA* and *FCRL3,* and adjusted p-value <0.05. All genes differentially expressed meeting log-fold change >1 are displayed in Supplementary Table 3.

Differential gene expression analysis comparing SC1 to all other B cells revealed a distinctive transcriptomic signature, including *CD38* and *CD9,* supporting classification as a transitional B cell population (Fig 6d), as well as many other genes not typically expressed at high levels in naïve B cells, including *TNFRSF18* (aka *GITR), PLD4, NEIL1,* and *ITM2C* (Fig 6e & Supplementary Table 3). SC1 displayed intermediate levels of ISG expression and notable down-regulation of several regulatory genes, including *ENTPD1*, recapitulating differential protein results of CD39 down-regulation, *FCRL3* and *IL10RA* (Fig 6e). SC2 was embedded between the transitional B cells and naïve B cells and was characterized by high ISG expression and intermediate levels of *CD38*, suggesting this population could also be a transitional population or a naïve B cell population in an altered IFN state, which we termed “IFN-hi naïve B” (Fig 6e). Transcriptomic expression of *ENTPD1* was not significantly different between IFN-hi naïve B and naïve B suggesting surface level expression of CD39 may be regulated post-transcriptionally in these naïve B cell populations. Together, the expansion of naïve B cells in the treatment-naïve state and the distinctive transcriptomic and proteomic properties of these naïve B cell subclusters, suggest a role for the B cell compartment in JDM.

## DISCUSSION

This is the first comprehensive evaluation of the immunophenotypes associated with disease activity in the peripheral blood compartment in JDM using multi-modal single-cell sequencing. We identified changes in cellular composition, characterized cell-specific IFN responses, and identified unique cell immunophenotypes associated with treatment-naïve disease. We also show that these changes occur both within individuals and between individuals with JDM by including longitudinal samples. Furthermore, by measuring both gene expression and surface protein expression, we were able to identify multi-modal features associated with TN-JDM. Cell surface proteins are not only important for cell phenotyping but also for carrying out key cellular functions providing greater insight into cellular behavior. By including oligonucleotide-barcoded antibodies for 50 cell surface proteins, we identified surface markers, such as CD56 in monocytes and CD39 in B cells, not typically associated with these immune cell types that displayed differential expression associated with disease activity in JDM. These results suggest that the peripheral blood compartment holds promise to help elucidate disease mechanisms and to identify candidate biomarkers and therapeutic targets.

In accordance with the striking transcriptomic IFN signature previously described in JDM blood and target tissues (3–6), as expected, we also identified a very strong IFN signature in our study. However, we did not anticipate that nearly every immune cell type would over-express a subset of IFN genes in the treatment-naïve state in our patient cohort. This pan-cell IFN rewiring strongly suggests that IFN signaling, either due to overactivation or lack of regulation, is a hallmark of JDM pathophysiology. CD16+ monocytes displayed the highest IFN expression, and other myeloid cell types (CD14+ monocytes and cDCs) and cytotoxic cell types (CD8+ T effector and CD56dim NK) also displayed higher IFN gene scores than other cell types suggesting these are the primary mediators of the IFN response observed in JDM. Using single-cell sequencing, we were able to tease apart IFN-related genes expressed across bulk cells from genes expressed in a cell-type specific manner overcoming limitations of prior bulk sequencing experiments where expression is confounded by cell composition. The combination of the pan-cell and cell-specific IFN responses we observed in JDM may explain why studies correlating IFN gene signatures and disease activity have been mixed (3, 7). The pan-cell IFN signature we identified was correlated with individual disease activity measures in our small study in bulk cells and more strongly in CD8+ effector T and CD56dim NK cells. Larger studies will be needed to determine the utility of this signature as a disease activity biomarker. This data, as well as prior literature (21, 22) would support the hypothesis that a carefully curated IFN gene signature correlates with disease activity in JDM. The IFN gene modules identified in our studies showed enrichment of both type I and II IFN signaling suggesting both pathways are activated in JDM, however, the involvement of type II IFN signaling in myositis continues to be controversial (4,23–25). Unfortunately, this data does not allow us further insight into the types of IFNs working on specific cell types or in specific tissues, and further highlights the complexity of IFN signaling in autoimmune diseases.

Our data have identified the peripheral blood monocyte compartment as a potential key player in JDM. In addition to exhibiting the strongest IFN response in TN subjects of all immune cell types and the greatest number of DEGs, CD16+ monocytes were quantitatively reduced in TN-JDM subject PBMCs and there was a trend toward reduction of CD14+ monocytes. We hypothesize this is due to tissue homing based on previous work from our group which identified strong enrichment of myeloid-derived cell types in muscle and skin microarray datasets (6) and recent cutting-edge work using mass cytometry imaging, which identified myeloid-derived cell types as the most abundant cell types in DM skin lesions (26). The role of CD16+ nonclassical monocytes in health and disease has been both controversial and context-dependent, but this cell type has traditionally been thought to be immune-regulatory (27). Prior work suggests CD16+ monocytes are predisposed to become migratory dendritic cells (28) or M2 anti-inflammatory macrophages (29). However, we identified a cluster of cells from TN-JDM subjects that were skewed toward an inflammatory and antigen-presenting phenotype more suggestive of pro-inflammatory M1 macrophages, suggesting this population is highly dysregulated in JDM. This CD16+ inflammatory population also displayed down-regulation of CD16, the Fc receptor FcγRIIIA. This receptor is internalized upon the binding of immune complexes (30) leading us to hypothesize that active signaling through this receptor may be the mechanism of CD16 downregulation. This model would provide a possible link between autoreactive B cells, myositis-specific antibodies, and activation of CD16+ inflammatory monocytes, which subsequently migrate to tissues and differentiate into pro-inflammatory macrophages sustaining local inflammation. This model is also supported by evidence that suggests IVIG, an effective therapy in JDM, may work through blockade of activating Fc receptors (30). Alternatively, these cell types may be highly plastic and recruited to tissues to support tissue repair, or they may be detected in peripheral blood because they lack the appropriate tissue homing receptors. Further work to determine the trajectory of these cell types in target tissues is needed to determine the utility of targeting this cell type therapeutically.

There is also growing evidence to support a role for myeloid cell types, especially CD16+ monocytes, in other autoimmune diseases, including in systemic lupus erythematosus (SLE) (31), another IFN-mediated disease closely related to JDM. In TN-JDM, we observed a skewing of both CD16+ and CD14+ monocytes toward an inflammatory and antigen-presenting state, co-expressing IFN and IL-1 axis genes, consistent with a finding recently described in childhood lupus (32). This study also identified a correlation with the lupus IFN signature in CD16+ monocytes. A similar inflammatory CD16+ non-classical monocyte population has also been identified in adult SLE peripheral blood (31). These findings also extend into tissue where single cell sequencing of infiltrating immune cells in the kidneys of lupus nephritis patients also identified a continuum of CD16+ macrophage cell types, including inflammatory CD16+ macrophages without a CD14 counterpart (33). In adult dermatomyositis, peripheral blood monocytes display increased expression of TLR’s (34). In our data, we also observed an increase in the number of inflammatory monocytes and the expression of inflammatory and IFN genes in one 6 month sample from a subject who was diagnosed with disease flare approximately three months later, a finding we are eager to follow up on as a potential subclinical marker of disease flare.

Different patterns of integrin and adhesion molecule surface expression distinguish monocyte populations in our study. CD11b and CD11c are well known markers distinguishing monocyte subsets, but we additionally observed CD49d as an integrin distinguishing CD16+ inflammatory monocytes from CD14+ inflammatory monocytes. There also appears to be a qualitative increase in CD49d in CD16+ inflammatory monocytes compared to non-inflammatory CD16+ monocytes, but we may have lacked power to detect a statistically significant difference for this marker between these two populations. CD49d is the alpha chain of the very late antigen 4 (VLA-4) integrin that binds to ligands fibronectin and VCAM-1 expressed on endothelium, the latter which was found to be up-regulated on muscle vessels in DM but not JDM (35). In Duchenne muscular dystrophy, a genetic myopathy with an inflammatory component, CD49d-expressing T cells displayed greater migratory responses and adhesion to myotubules, and inhibition of CD49d is being studied as a novel therapeutic option (36).

We additionally see a striking expression of CD56 on CD14+ monocytes that is quantitatively reduced in TN patients and increases with treatment. CD56-expressing monocytes have also been identified in RA (37) and Crohn’s disease (38) where they were found to be expanded; however, in RA the number of these cell types decreased with treatment, whereas we see the opposite effect in JDM patients for unknown reasons. We also identified the same directional change in expression of CD56 in CD8+ effector T cells with increased expression in inactive JDM compared to TN-JDM. CD56-expressing monocytes have also been described to be expanded in many cancers and identified as CD56+ tumor-infiltrating dendritic cells with the capacity to lyse tumor cells in the presence of IFN-⍺, suggesting they are important in the anti-tumor response (39). Likewise, CD56-expressing CD8+ T cells display increased cytotoxicity (40), and expression of CD56 by CAR-T cells improves their survival and anti-tumor response (41). One possible explanation is that CD56-expressing monocytes and CD8+ T cells display increased cytotoxic behavior and are increased in tissues, thus decreased in peripheral blood in the treatment-naïve state, but it is also possible that this receptor plays an important immunoregulatory role in JDM that is deficient at disease onset.

We identified expansion of a transitional B cell population with increased CD24 and CD5 expression as well as CD39 down-regulation and differential expression of several immunoregulatory genes, including up-regulation of *TNFRSF18 (GITR), PLD4*, and down-regulation of *ENTPD1, IL10RA*, and *FCRL3*. It is unclear whether this transitional population is a precursor to naïve B cells or an altered naïve B cell state in response to ISG stimulation. In this case, the ISG-hi naïve B cell population may represent a continuum of naïve B cell activation. Alternatively, these may represent two separate populations: one IFN-stimulated naïve B cell population and one transitional B cell population with distinct functional properties. Prior literature also identified an expansion of CD19+CD24hi, CD38hi transitional B cells in JDM and a correlation of the proportion of these cells with the type I IFN signature in B cells (12). It seems plausible that the transitional B cell population identified in this study is analogous providing further support that this cell type may be relevant to the disease process. We identify several additional features of this immunophenotype, including key genes and surface proteins, like CD5 expression.

The role of CD5 in naïve B cell populations has been controversial, described by some as an activation marker and by others as a distinct phenotypic B cell subset marker (42). In SLE, a CD5+ pre-naïve B cell population has been described that displayed functional properties of plasma cell differentiation and antigen-presentation, which the authors postulated to represent a mechanism of autoreactive B cell escape (43). We hypothesize that B cell development is altered due to the overactive IFN response in JDM, which may result in activation and proliferation of these transitional B cell populations, some of which may escape tolerance checkpoints. Furthermore, we observed, in both naïve and memory B cells, down-regulation of CD39 in TN-JDM and increased expression during the first 6 months of treatment during which all patients demonstrated a decrease in disease activity. The down-regulation of CD39 in these naïve B cell populations in TN patients may also contribute to such potential escape of autoreactive B cells from regulatory mechanisms like CD39-mediated immune regulation, a phenomenon also described in rheumatoid arthritis (44), or IL-10 production (12). CD39, or *ENTPD1*, contributes to adenosine production which has several anti-inflammatory effects. The role of this receptor is most well characterized in Tregs in human autoimmune disease, including SLE and RA, and is also of interest as a therapeutic target (45).

The expression of *TNFRSF18 (GITR)*, a co-stimulatory molecule, in JDM transitional B cells is of interest. It may simply represent a marker of transitional B cells with no functional consequences on B cell maturation, as suggested in murine studies (46), or represent an important signaling pathway by which these cells become dysregulated or influence lymphocytes. Expression of GITR ligand by B cells has been found to play an important role in the maintenance of Treg populations in mouse autoimmune models, but there is little to no data studying the role of GITR in transitional B cells in human disease states. Given the success of many immunotherapy agents, there is great interest in this receptor as a therapeutic target for both cancers and autoimmune diseases. Likewise, *PLD4* is also of interests as it is an exonuclease with regulatory role in breaking down nucleic acids, and genome-wide genetic variants have been associated with SLE, systemic sclerosis and RA (45). *PLD4*-deficient mice develop a severe inflammatory phenotype emphasizing the importance of this gene in immune regulation (47). Functional studies of JDM B cell populations and identified gene pathways in a disease-specific context as well as assessment of the clonality and antigen specificity of B-cell receptors will be important next steps in determining the role of transitional B cells in JDM and relationship to the myositis-specific autoantibodies observed in JDM.

There are important limitations to consider when interpreting the results of this study. While this study is strengthened by the inclusion of JDM treatment-naïve samples with longitudinal time points, we did not have healthy controls samples to be able to determine the specificity of the cell-specific signatures and cell types we identified to JDM. Furthermore, due to the rarity of this disease, our sample size was limited, and this did influence our ability to test the association between cell type composition and disease activity with adequate power. Larger studies to test the association between the number of circulating naïve B cells and CD16+ monocytes, or, perhaps, the ratio of these cell types with disease activity, will be important to test in larger cohorts, and additional alterations in immune cell type composition may emerge. Our analysis was also limited to the measurement of 50 pre-specified cell epitopes, which does not fully reflect the compendium of cell surface antibodies, though is the largest number of cell epitopes measured simultaneously to date in JDM. Lastly, JDM is a heterogeneous disease, so it is also possible that certain clinical phenotypes are associated with distinct cell immunophenotypes or subtle signaling pathways not detectable in small studies. Certainly, larger single cell studies considering disease heterogeneity, such as MSA subtype, will be needed in the future to understand if there is an immunologic correlate to explain the disparate clinical phenotypes observed in JDM.

Our study demonstrates the strengths of these experimental and analytic methods to provide great insights into the immunology of autoimmune diseases as well as evidence and data that may inform future immunologic studies. This work is the first comprehensive multi-modal single-cell analysis of the peripheral blood compartment in JDM, and the data will be made available for other myositis researchers to investigate genes and proteins of interest in a cell-specific manner. We hope these findings and accompanying data will lead to many rich insights and hypothesis generation for future JDM research and ultimately help to inform precision medicine approaches to disease management to improve patient outcomes.

## METHODS

### Patients

This study was approved by the University of California, San Francisco (UCSF) Institutional Review Board. Subjects meeting Bohan and Peter criteria for JDM (48), modified to include MRI as a possible diagnostic modality reflective of current practice, were recruited from the UCSF Pediatric Rheumatology Clinics in San Francisco and Oakland. All subjects provided informed consent, as well as informed assent, when age-appropriate. Patients could be enrolled at any time in their disease course. At each study visit, patient clinical data, which included items contained in the IMACS core consensus data set (49), was collected in a secure REDCap database. Patients enrolled at diagnosis also had follow up study visits at approximately 2, 4 and 6 months into treatment. For this study, inactive disease was defined using a modified PRINTO criteria for inactive disease (50): creatinine kinase <=150, manual muscle testing (MMT)-8<=78, and physician global visual analog score (VAS) <=0.5 as well as clinical judgement of inactive disease. We used a threshold of <=0.5 rather than 0.2 because our data collection forms included checked boxes in increments of 0.5 for VAS scores, and we did not collect CMAS measurements. Demographics, disease characteristics, and median disease activity measures for each patient group are summarized in Table 1. These measures are displayed graphically in Supplementary Figure 1.

### Sample Processing & Multi-modal Single Cell Sequencing

Peripheral blood samples were collected at each study visit and processed by the Pediatric Clinical Research Core Sample Processing Lab. Peripheral blood mononuclear cells (PBMCs) were collected in SepMate tubes (n=4) or CPT tubes (n=16), isolated per each manufacturer’s guidelines, and cryopreserved in liquid nitrogen. Our experimental protocol followed the manufacturer’s user guide (10X 3’V3 Document CG000185 Rev B, 10X Genomics) with certain modifications to isolate and amplify antibody-derived DNA tags (ADTs). Note these experiments were carried out using early access kits from BD Genomics before the implementation of commercially-available single-cell protein/RNA assays (e.g. Feature Barcoding, 10x Genomics; BD Abseq, BD Genomics, Supplementary Table 4), and researchers are recommended to use those newer solutions for any follow-up studies as the techniques and reagents have been refined. For the experiment, PBMCs from 20 distinct samples were gently thawed in a 37°C water bath and re-suspended using a pipette set to 1 mL. Cell counts and viability were determined using a Cellometer Vision (Nexcelcom) with AOPI staining (Nexcelcom cat. CS2-0106-5ML). Cells were multiplexed into four pools with five samples each (2×10^5^ cells/sample) into separate 5 ml tubes (Falcon cat. 342235). Longitudinal samples from the same individual were aliquoted to distinct pools to enable genetic demultiplexing and experimental time points were also mixed within each well to avoid confounding time-related batch effects. After pooling, cells were resuspended in 90 μl of 1% BSA in PBS and Fc blocked with 10 μl Human Trustain FcX (Biolegend cat. 422302) for 10 minutes on ice then stained on ice for 45 minutes with a pool of 50 antibodies in 100 μl, for a final staining volume of 200 μl. Antibodies were pooled on ice with 2.2 μl per antibody per 1×10^6^ cells (BD Genomics). Cells were quenched with 2 ml 1% BSA in PBS and spun at 350xg for 5 minutes and further washed two more times with 2 ml of 1% BSA in PBS. After the final wash, cells were resuspended in 100 ul and strained through a 40 μM filter (SP Bel-Art cat. H13680-0040). Each pool was loaded and processed into a respective well (4 wells total, 4×10^4^ cells/well) as in the manufacturer’s protocol. The 10x instrument was run and post-GEM RT and cleanup were done as according to manufacturer’s protocol. Starting at cDNA amplification, modifications to the protocol were made: 1 μl of 2 μM additive primer (BD Genomics, beta kit) specific to the antibodies tags was added to the amplification mixture. During the 0.6X SPRIselect (Beckman Coulter, B23318) isolation of the post-cDNA amplification reaction cleanup, the supernatant fraction was retained for ADT library generation. Subsequent library preparation of the cDNA SPRI-select pellet was done exactly according to protocol, using unique SI PCR Primers (10x Genomics). For the ADT supernatant fraction, a 1.8X SPRI was done to isolate ADTs from other non-specifically amplified sequences, followed by a sample index PCR. Sample index PCR for the ADTs was done using the cycling conditions as outlined in the standard protocol (15 cycles) but using different unique SI-PCR Primers such that all libraries could be mixed and sequenced together. Subsequent SPRI selection was performed, and all libraries were quantified and analyzed via Qubit 2.0 (Fisher) and Bioanalyzer (Agilent), respectively, for quality control. Libraries were mixed and sequenced on 1 lane of a NovaSeq S4 using the recommended number of cycles.

### Alignment, demultiplexing, and doublet removal

CellRanger (v3.1.0) was run to align reads to the GRCh38 genome build and generate counts matrices and binary alignment files (BAMs). Each BAM was inputted into freemuxlet for donor-of-origin annotation and doublet removal. To assign cells to donors of origin in our multiplexed design, we used the genetic demultiplexing tool freemuxlet and a sample matching script, each being part of the popscle suite of population genetics tools (https://github.com/statgen/popscle) to assign cells to donors as previously described (51). To create the external genotype reference, DNA was extracted from whole blood samples for each patient and genotyped using the OmniExpressExome array at the UC Berkeley Vincent J. Coates Genomics Sequencing Laboratory. Freemuxlet and the sample matching script were run, yielding a 1 to 1 mapping of droplet barcode clusters to individuals. Heterotypic doublets detected by demuxlet were removed, and additional homotypic doublets (i.e. two cells from the same individual co-encapsulated) were removed using doubletdetector (52).

### Quality control and processing

Gene and protein expression matrices (4 each) were subsequently processed using Scanpy (v1.5.1) (53). To identify and filter low quality cells, we visualized the log-normalized distributions of mRNA counts, protein counts, and gene counts for each of the four wells and filtered counts at the tails of these distributions. The percent mitochondrial gene expression for each well displayed a similar distribution, so we applied the same cutoff of filtering cells with >15% mitochondrial gene expression to all four wells. After filtering and doublet removal, we analyzed 55,564 cells. The average number of cells sequenced per sample was 2,778. Subject N-4 had significantly few cells sequenced than the three other newly diagnosed patients (n=2,663), compared to N-1 (n=13,812), N-2 (n=11,985), N-3 (n=11,633)), particularly for visits 2 and 3 where only 12 and 116 cells were recovered, respectfully. This was taken into account for all downstream analyses. A similar number of cells, ∼14,000 was recovered from each of the four 10X wells.

The four mRNA matrices were then concatenated and log-normalized. We then extracted a set of highly-variable genes using the Scanpy function “highly_variable_genes” and then selecting a set of genes with high mean expression and dispersion. The protein matrices were also concatenated and log-normalized, and the both RNA and protein matrices were merged for down-stream join clustering. The Scanpy commands “regress_out”, to control for the effect of mitochondria gene count and total mRNA count, and “combat” (54), to control the batch effect of the four wells, were used. Batch correction using ComBat was able to correct for technical replicates except in CD14+ monocytes, where there appears to be some residual sample-specific effects especially related to cluster 18. These effects could be due to batch or other covariates such as sex, which is consistent with previous literature showing significant inter-individual variation in monocytes (55, 56) (Supplementary Fig. 4). We accounted for these effects by analyzing these clusters collectively in down-stream analysese. Clusters were otherwise independent of patient effect, however, there was some patient-level heterogeneity: N-2 cells make up a large majority of CD8+ effector T cells and N-1 and N-3, both males, appear to cluster together within the CD14+ monocyte population (Supplementary Fig. 5). Principal component analysis was applied to identify 30 PCs, and the “neighbors” command was used to compute a neighborhood graph using the default size of 15. We then embedded the neighborhood graph using Uniform Manifold Approximation and Projection (UMAP) (57) for subsequent visualization using the default settings.

### Clustering and cell type annotation

Clustering was then performed using the set of highly variable genes and all proteins and applying the Leiden algorithm (58) with a resolution of 1.5, which identified 24 cell clusters, 23 of which were immune cell clusters (cluster 22 was platelets), including all major immune cell populations (Supplementary Fig 2, Fig. 2a). Clusters were manually annotated by visualizing canonical protein surface markers on the UMAP and by calculating marker genes and proteins of each cluster. Two of these clusters, 13 and 15, could not be annotated and expressed both CD8 and mRNA transcripts for the gamma and delta chains of the T cell receptor. These two clusters were further subclustered at a low resolution using the “restrict_to” parameter in the Leiden function in Scanpy to identify distinct CD8+ memory T cell and gamma-delta T cell populations. Cells were annotated using canonical RNA and cell surface markers (Fig. 2a & Supplementary Fig. 3).

### Cell Type Compositional Analysis

To determine cell types enriched in active disease, cell type proportion within newly diagnosed patients was compared between visit 1 (V1), or treatment-naïve (TN) state, and visit 4 (V4) using a paired t-test and significance threshold of p<0.05. All four patients exhibited a decline in disease activity levels during the first 6 months of treatment. We also compared cell type proportion between TN-JDM and inactive JDM (I-JDM) using a t-test and a significance threshold of p<0.05. For both analyses, percent composition was calculated and visualized with dittoSeq’s “dittoBarPlot” function (59).

### Differential gene and protein expression

To calculate cell type differential gene and protein expression between disease states, we used the DESeq2 package (v1.28.1) (60), using the inputs recommended for single-cell data in the package vignette (test = “LRT”, reduced = ∼1, sfType = “poscounts”, useT = TRUE, minmu = 1e-6, minReplicatesForReplace = Inf) and included batch as a covariate in the design formula. We used a log-fold change cutoff of >1 for differential gene expression or >0.5 for protein expression, and a false discovery rate <0.05. Only genes and proteins expressed in at least 10% of cells for each cell populations were assessed. Differential expression was not calculated for plasmablasts, due to low cell numbers, or platelets.

### Global cell-specific transcriptional signature

To identify and visualize the global cell-specific transcriptional signature, we collated the set of non Y-chromosome genes that were differentially expressed in at least one cell type in both analyses longitudinal and cross-sectional analyses. The psuedobulk mean expression per sample group, defined as visit 1, 2, 3, 4 and inactive, for each of the genes per cell type was calculated and visualized using dittoSeq’s ‘dittoHeatmap’ (59). Columns were ordered by cell type and group by increasing time from diagnosis, which also correlated with decreasing disease activity levels. Unsupervised hierarchical clustering by Euclidean distance was then applied to cluster the genes into distinct modules using k=10. Module scores were then calculated as the sum of mean expression of all module genes for each pseudobulk. Correlations of module scores (minimum of 10 cells per case) to X-metric was then calculated with using the Pearson correlation and visualized as dot & scatter plots using ggplot2 (61). Gene Ontology enrichment analysis

### Identifying Cell Type Subclusters

To identify subclusters within selected cell populations, a second round of clustering was applied using the “restrict_to” parameter in the Leiden function in Scanpy and a low clustering resolution of 0.2 for CD16+ monocytes and 0.3 for naïve B cells. The proportion of cells from each patient group was calculated and a chi-square test was applied to determine if the composition of subclusters differed. Differential gene expression and differential protein expression for each subcluster compared to the canonical cell population was calculated using DESeq2 in the same method as described above.

## Data Availability

The data associated with this study will be made available in the pertinent databases upon acceptance to a peer-reviewed journal.

## Author contributions

JN designed the research study, enrolled subjects and performed clinical phenotyping, analyzed data, and wrote the manuscript.

GH planned and conducted experiments, analyzed data, and wrote the manuscript.

DB analyzed data and wrote the manuscript.

YS planned and conducted experiments and revised the manuscript.

DL planned and conducted experiments and revised the manuscript.

SK designed the research study, enrolled subjects and performed clinical phenotyping, and revised the manuscript.

CJY designed the research study, analyzed data, and revised the manuscript.

MS designed the research study, analyzed data, and revised the manuscript.

## Acknowledgements

The authors would first like to express gratitude for the patients with JDM and their families who contributed their samples and data to this study. We would also like to acknowledge and provide thanks to the funding sources, core facilities, and individuals who supported our work: This work was supported by Grant 2019124 from the Doris Duke Charitable Foundation, grants from the Cure JM Foundation, a grant from PREMIER, a NIH/NIAMS P30 Center for the Advancement of Precision Medicine in Rheumatology at UCSF (P30AR070155), and a CARRA Arthritis Foundation grant. The authors wish to acknowledge CARRA and ongoing Arthritis Foundation financial support of CARRA. We would also like to acknowledge the Institute of Human Genetics Genomics Core at UCSF, Center for Advanced Technology at UCSF, and the UC Berkeley Vincent J. Coates Genomics Sequencing Laboratory. We thank the members of the UCSF Pediatric Rheumatology Division for helping to recruit patients: Emily von Scheven, William Bernal, Michael Waterfield, Erica Lawson, Alice Chan, Geraldina Lionetti, Nicole Ling, Tara Valcarcel, Andy Nguyen, William Soulsby, Julia Shalen, and Sara Haro, and our research coordinators, Bhupinder Nahal and Kathy Nguyen.

