## Supplementary for "Multi-modal single-cell sequencing identifies cellular immunophenotypes associated with juvenile dermatomyositis disease activity"

### Supplementary Figure 1: Disease Activity Measures for at each time point or sample group.

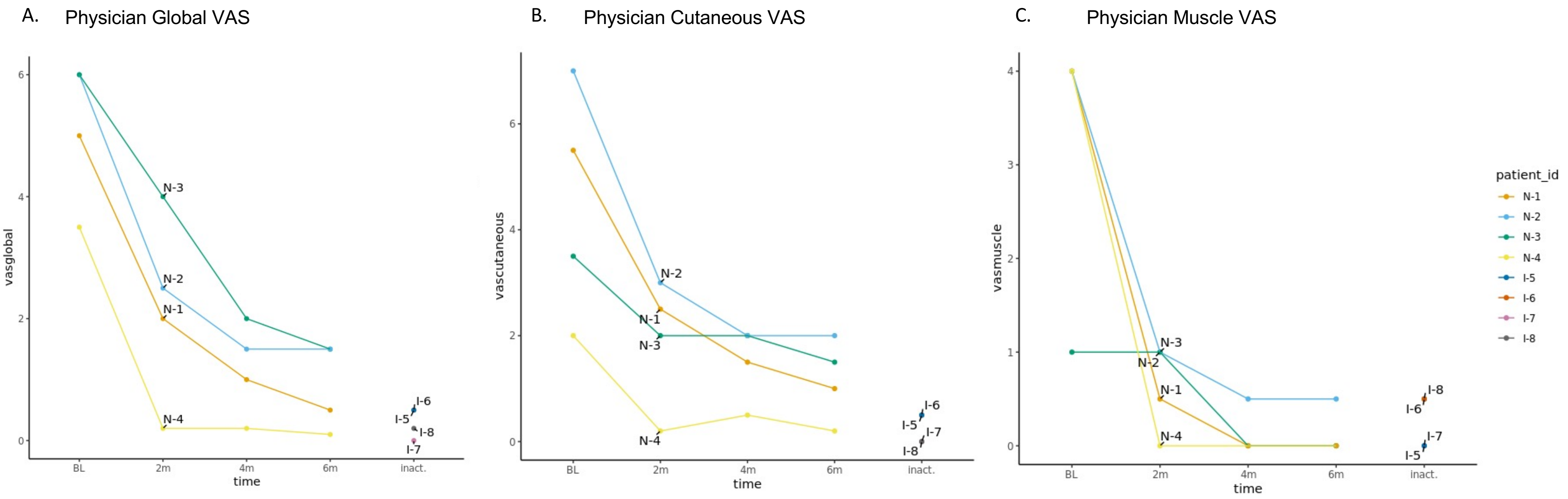


**Supplementary Fig. 1**: The x-axis represents represents longitudinal time points corresponding to visit 1 (TN state), visit 2 (~2 months), visit 3 (~4 months), visit 4 (~6 months) and the four inactive disease samples for A. Global, B. Cutaneous, and C. muscle physician VAS scores. VAS = visual analog scale.

### Supplementary Figure 2: Cell clusters identified by Leiden clustering

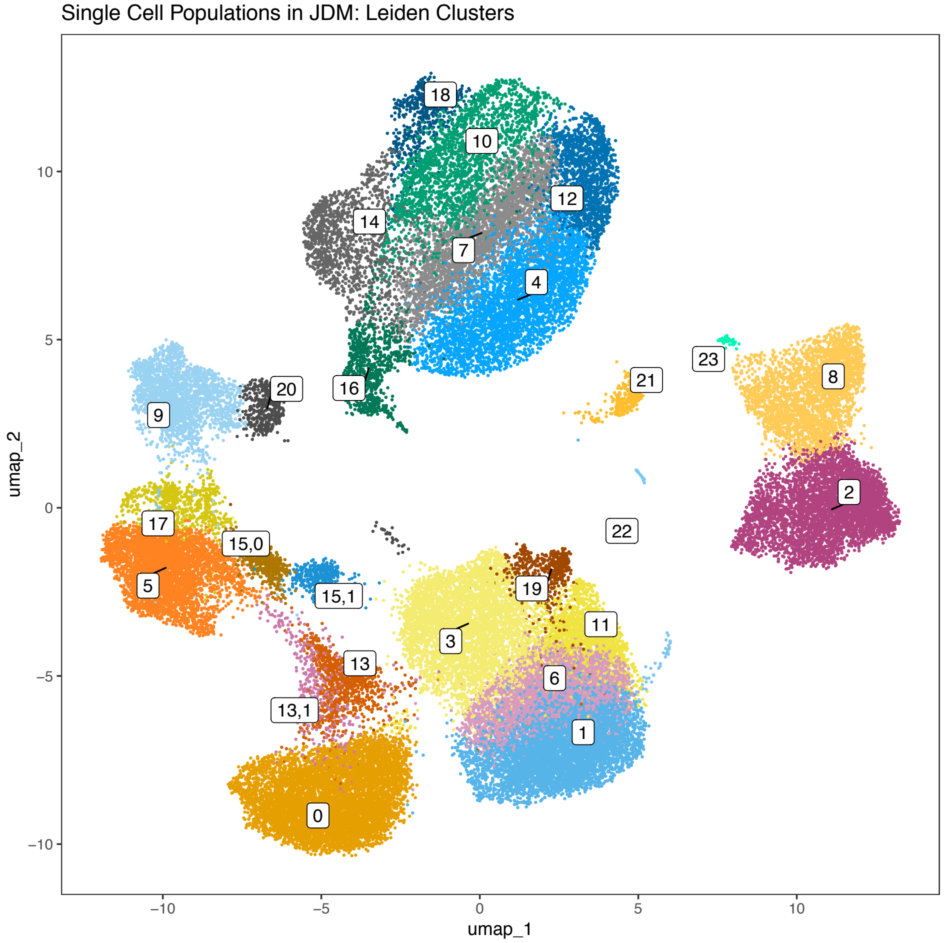


**Supplementary Fig. 2**: UMAP plot visualizing 23 cell clusters identified by Leiden clustering using a clustering resolution of 1.5.

### Supplementary Figure 3: Cluster Annotation Markers

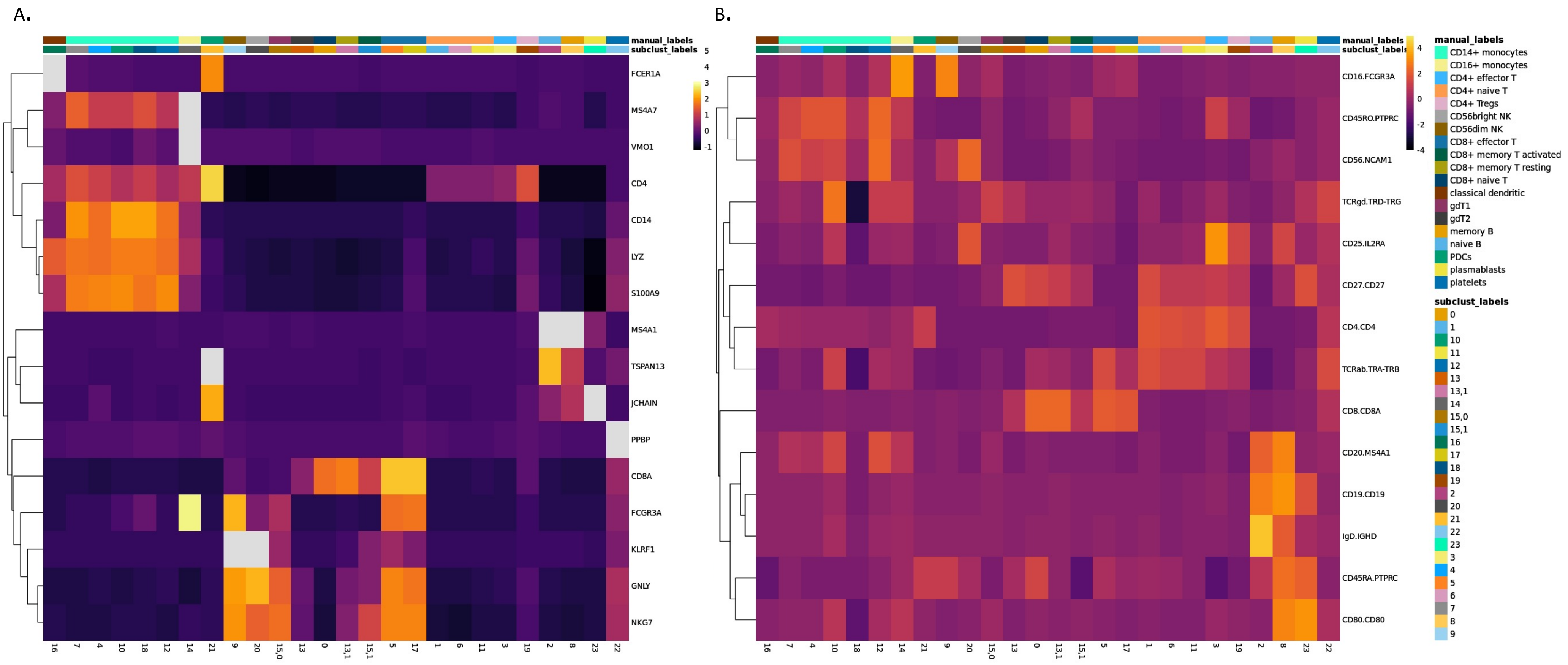


**Supplementary Fig. 3:** RNA (A) and ADT (B) markers used for annotating cell clusters visualized using hierarchical clustering of features. Columns are ordered by manual annotations.

### Supplementary Figure 4:  Batch effects

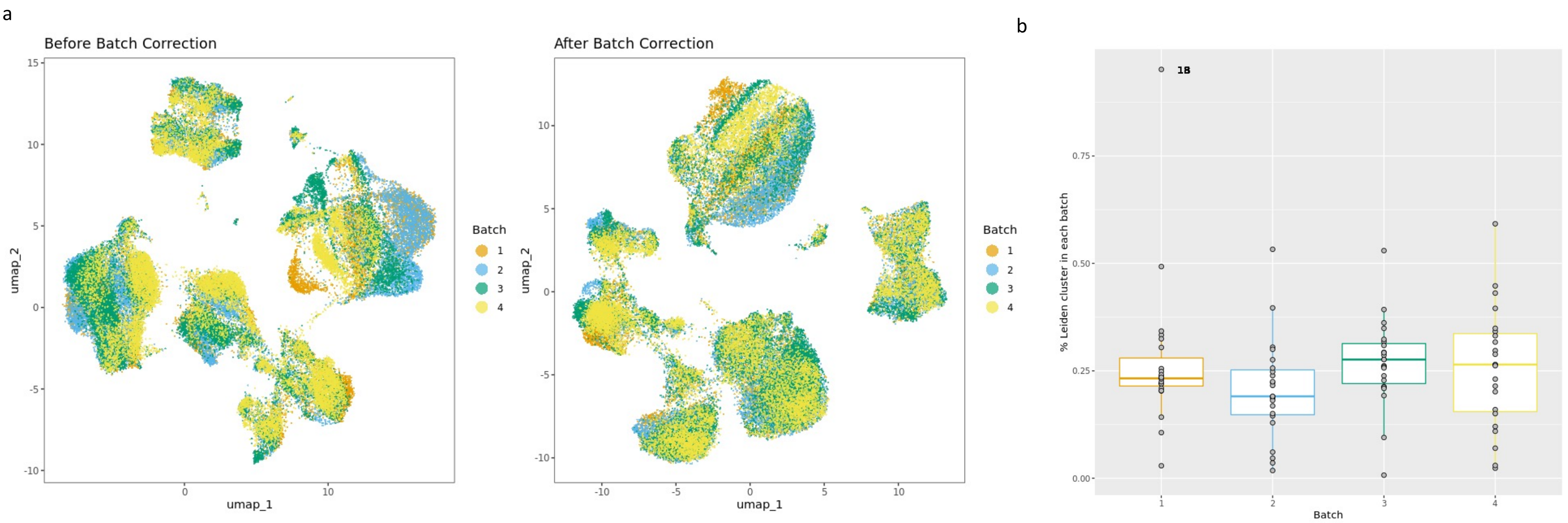


**Supplementary Fig. 4**:  A. UMAP plots before and after batch correction using ComBat to correct for the four 10X wells.  B. Percent of Leiden clusters from each batch.

### Supplementary Figure 5:  UMAP colored by subject

A.

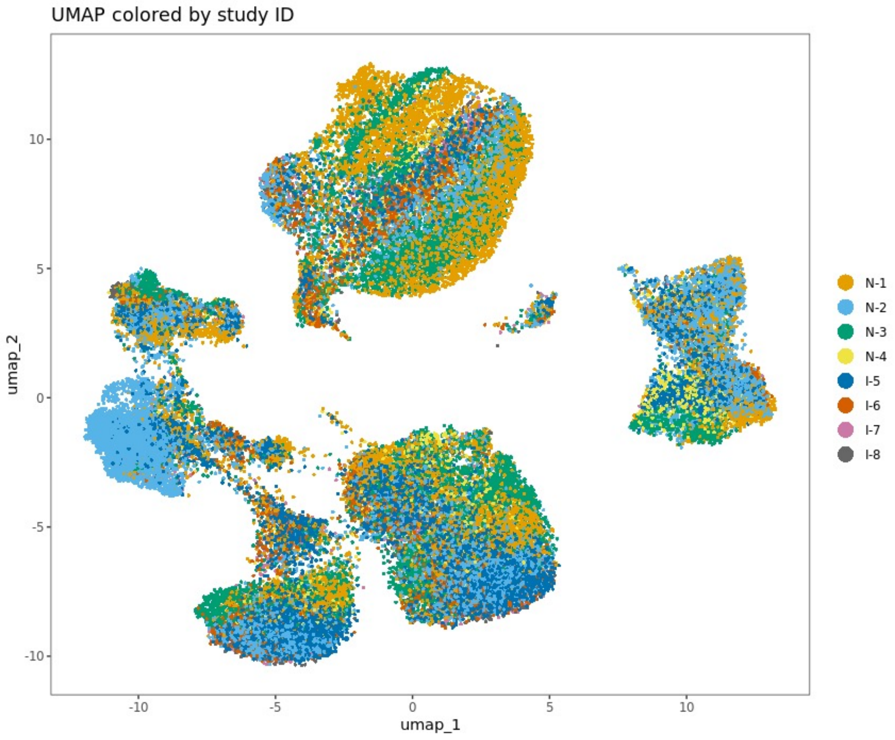


B.

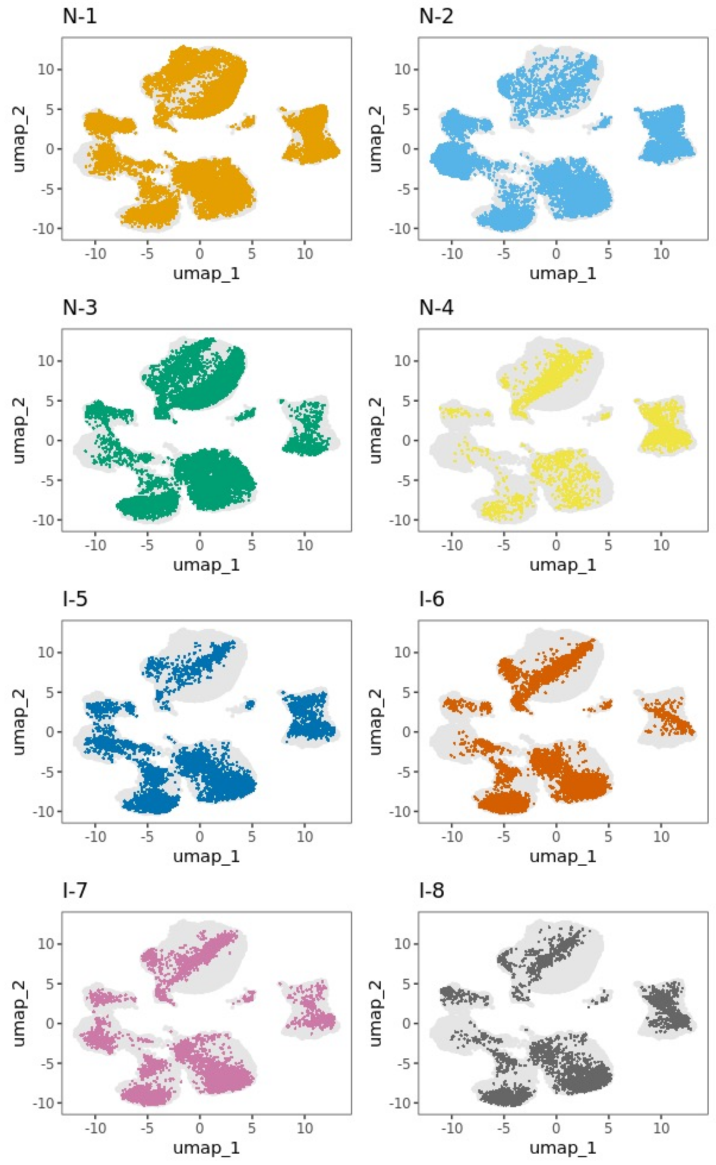


**Supplementary Fig. 5**: UMAP of all cells colored by study ID together in panel A and individually in panel B.

SF6

A.

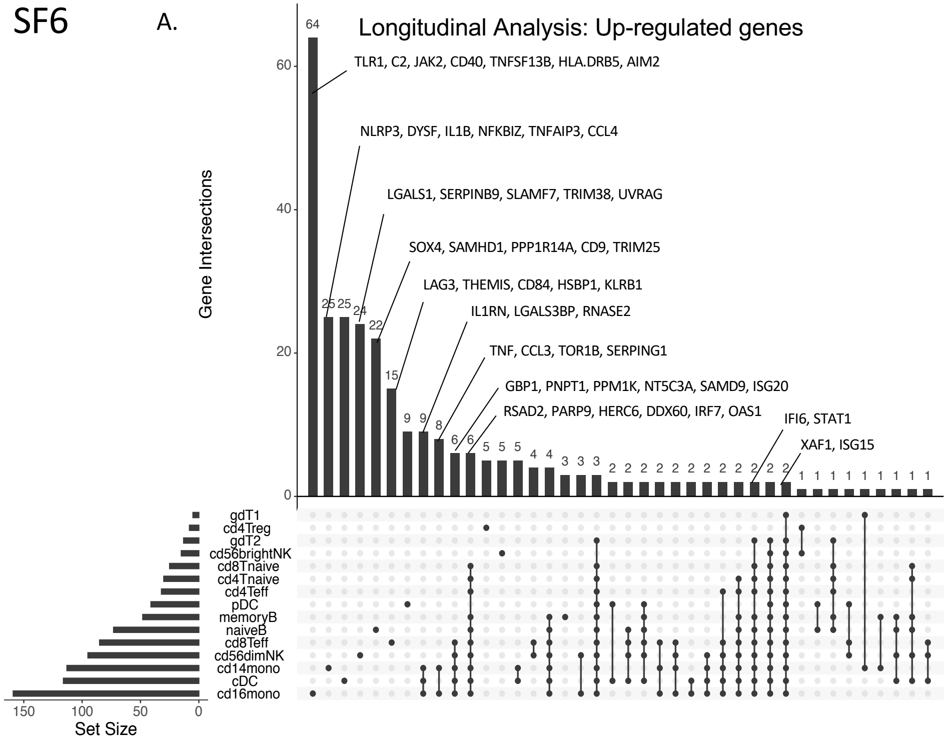


B.

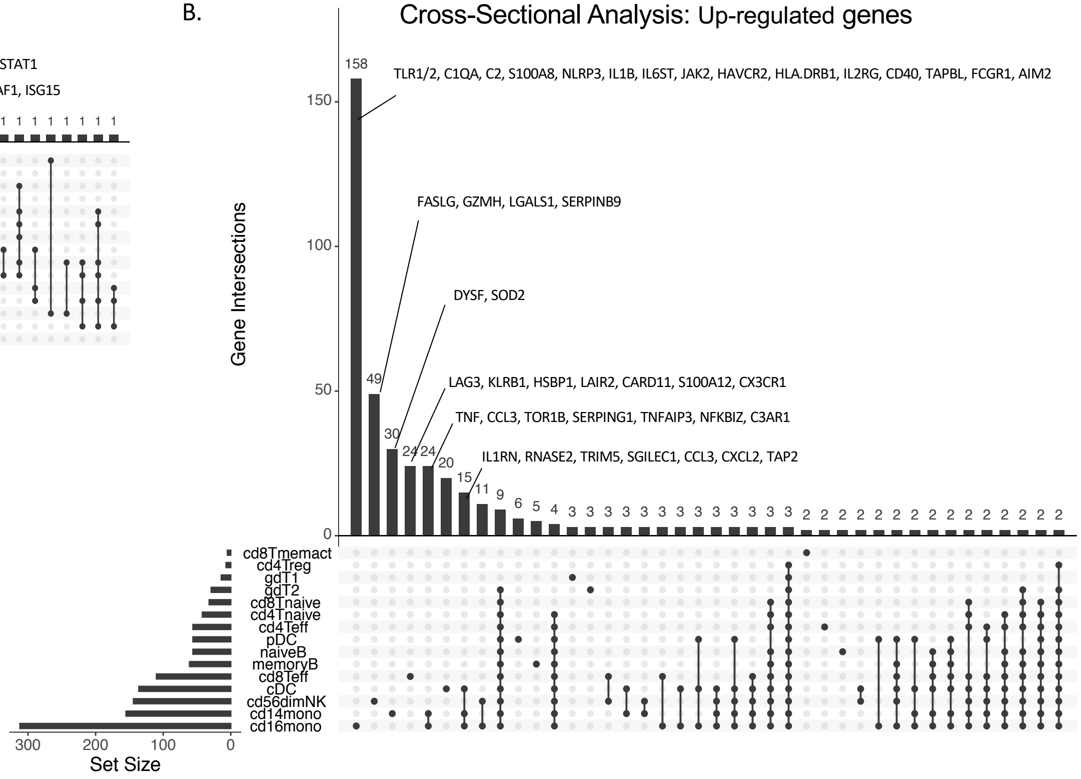


**Supplementary Fig. 6:** Upset plots displaying the overlap of differentially expressed genes (DEGs) up-regulated in each cell type for the longitudinal analysis (A) and cross-sectional analysis (B). Genes of interest are annotated manually. The horizontal bar graph shows the total number of DEGs for each cell type and the vertical bar graph shows the total number of genes that are either unique to a cell type (a single dot) or shared across multiple cell types (multiple dots connected in a line).

SF7

A.

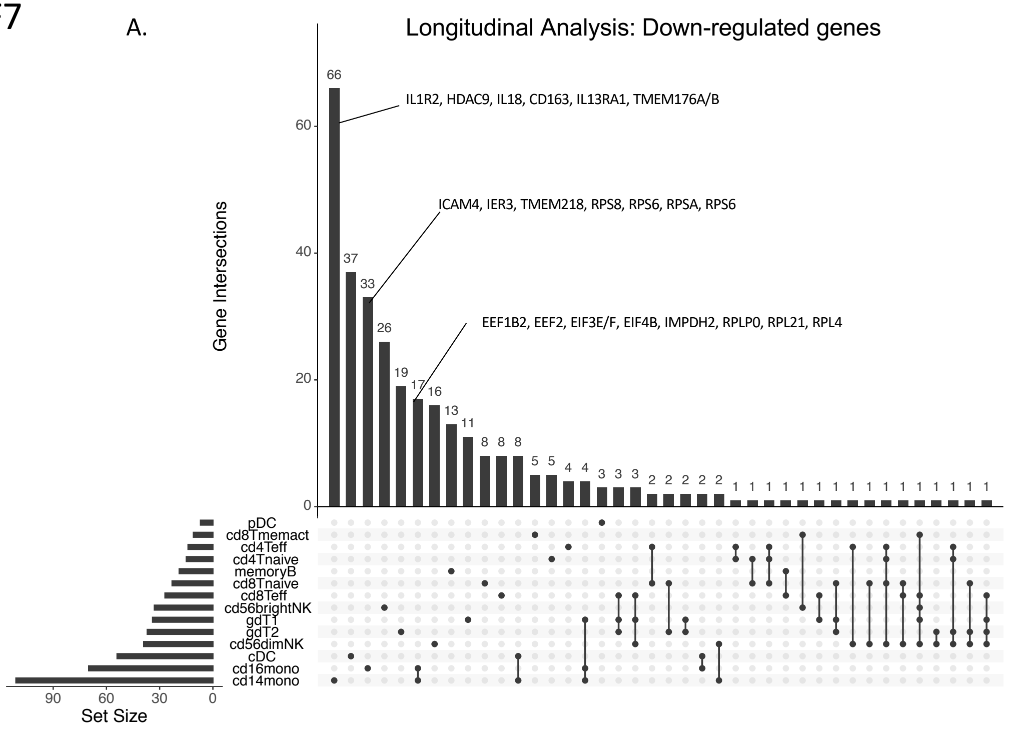


B.

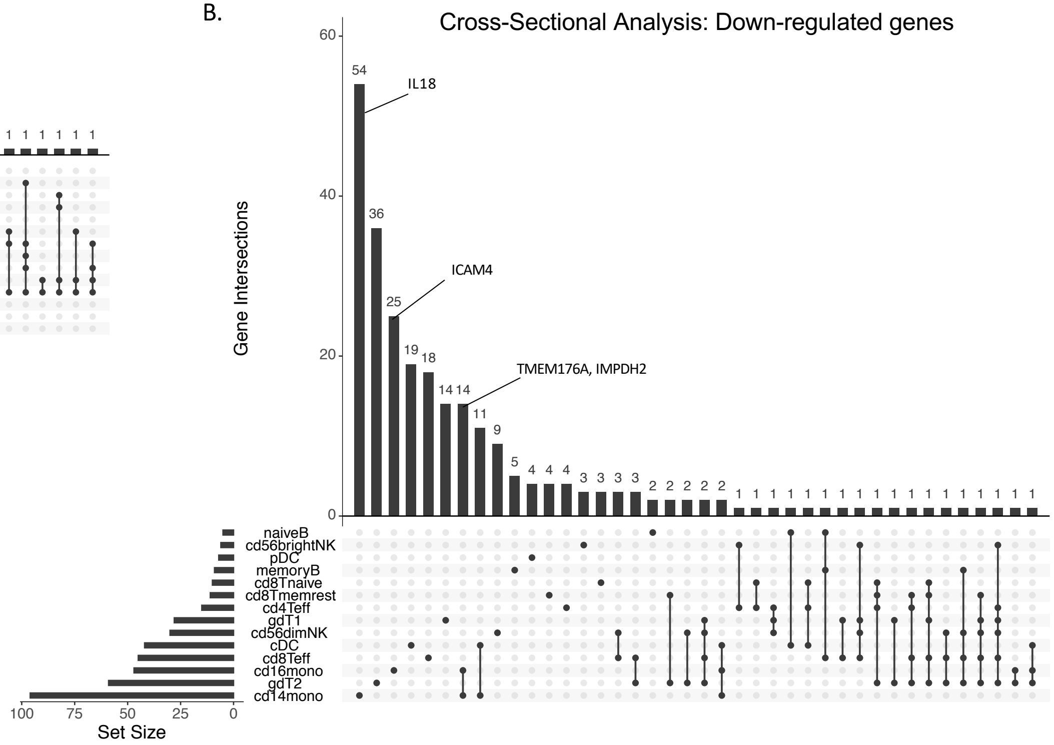


**Supplementary Fig. 7:** Upset plots displaying the overlap of differentially expressed genes (DEGs) down-regulated in each cell type for the longitudinal analysis (A) and cross-sectional analysis (B). Genes of interest are annotated manually. The horizontal bar graph shows the total number of DEGs for each cell type and the vertical bar graph shows the total number of genes that are either unique to a cell type (a single dot) or shared across multiple cell types (multiple dots connected in a line).

### Supplementary Figure 8

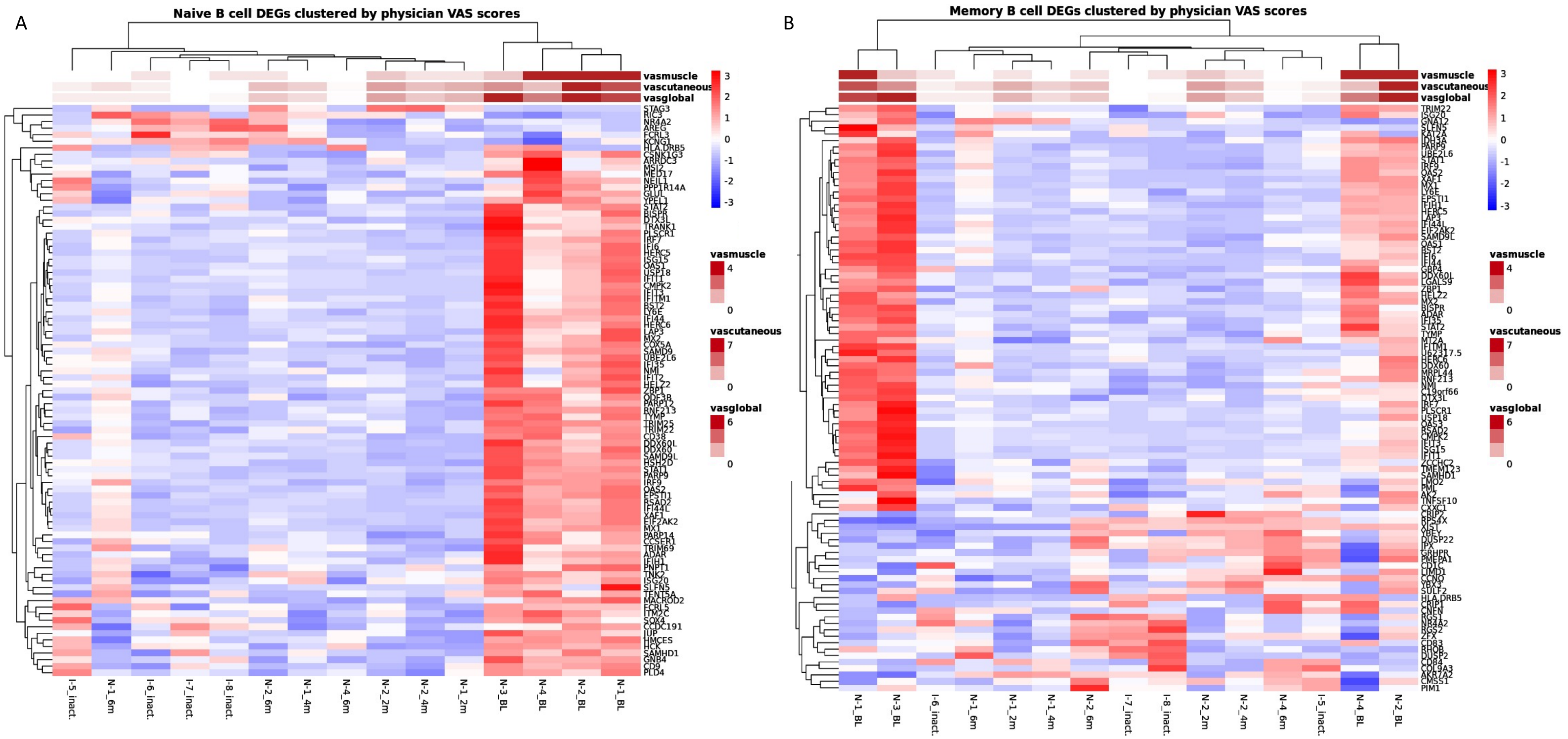


**Supplementary Fig. 8:** Heatmaps displaying hierarchical clustering of the psuedobulk expression of the union of genes differentially expressed in both longitudinal and cross-sectional analyses in naïve B cells (A) and memory B cells (B). Samples are annotated by disease activity measures including physical VAS scores for global ("vasglobal"), cutaneous ("vascutaneous"), and muscle ("vasmuscle") domains.

### Supplementary Figure 9

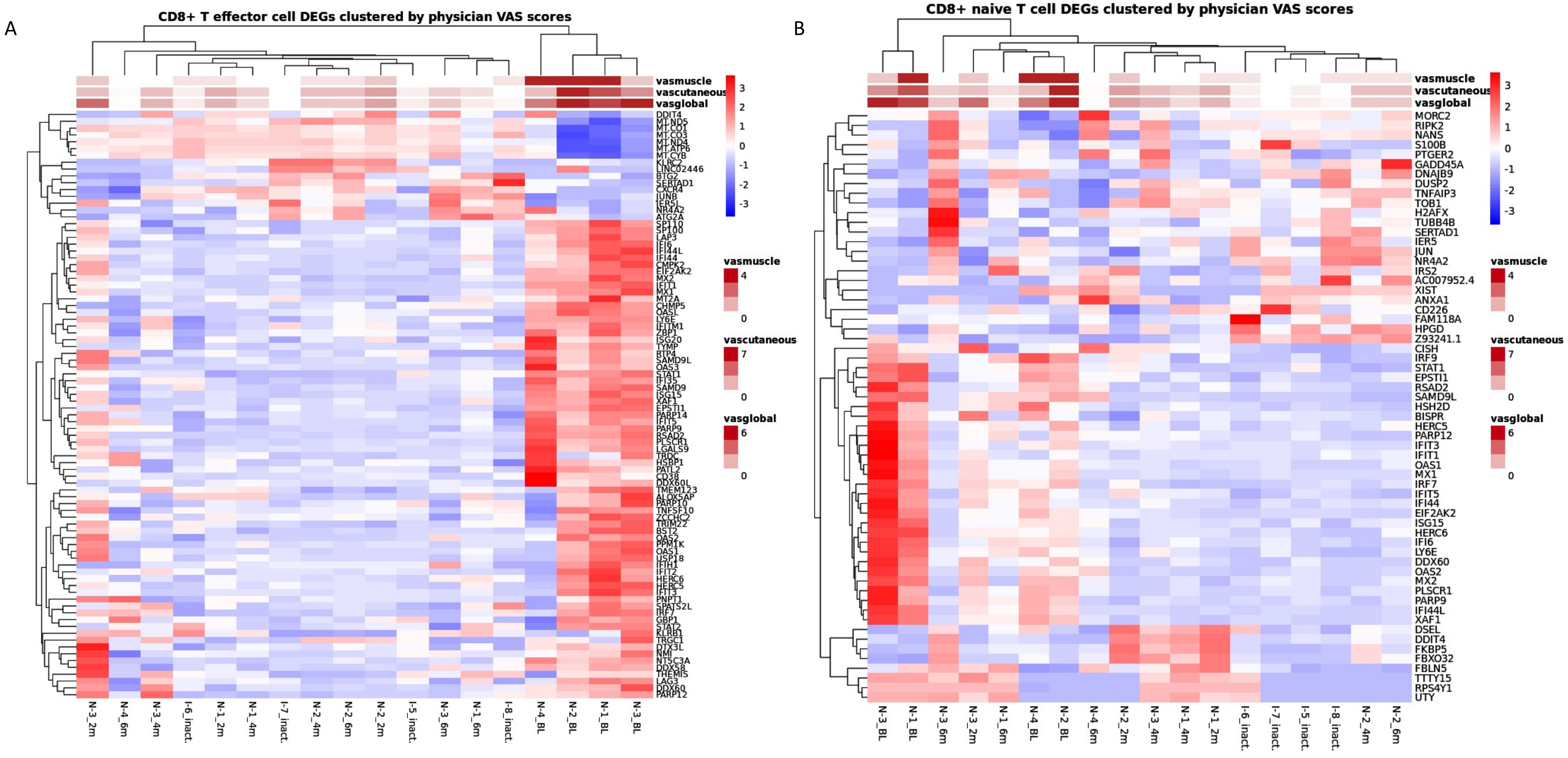


**Supplementary Fig. 9:** Heatmaps displaying hierarchical clustering of the psuedobulk expression of the genes differentially expressed in both longitudinal and cross-sectional analyses. The set of genes in CD8+ effector T cells is the intersection of genes expressed in both analyses (A) and the set of genes in CD8+ naïve T cells is the union of genes expressed in both analyses (B). Samples are annotated by disease activity measures including physical VAS scores for global ("vasglobal"), cutaneous ("vascutaneous"), and muscle ("vasmuscle") domains.

#### Supplementary Figure 10

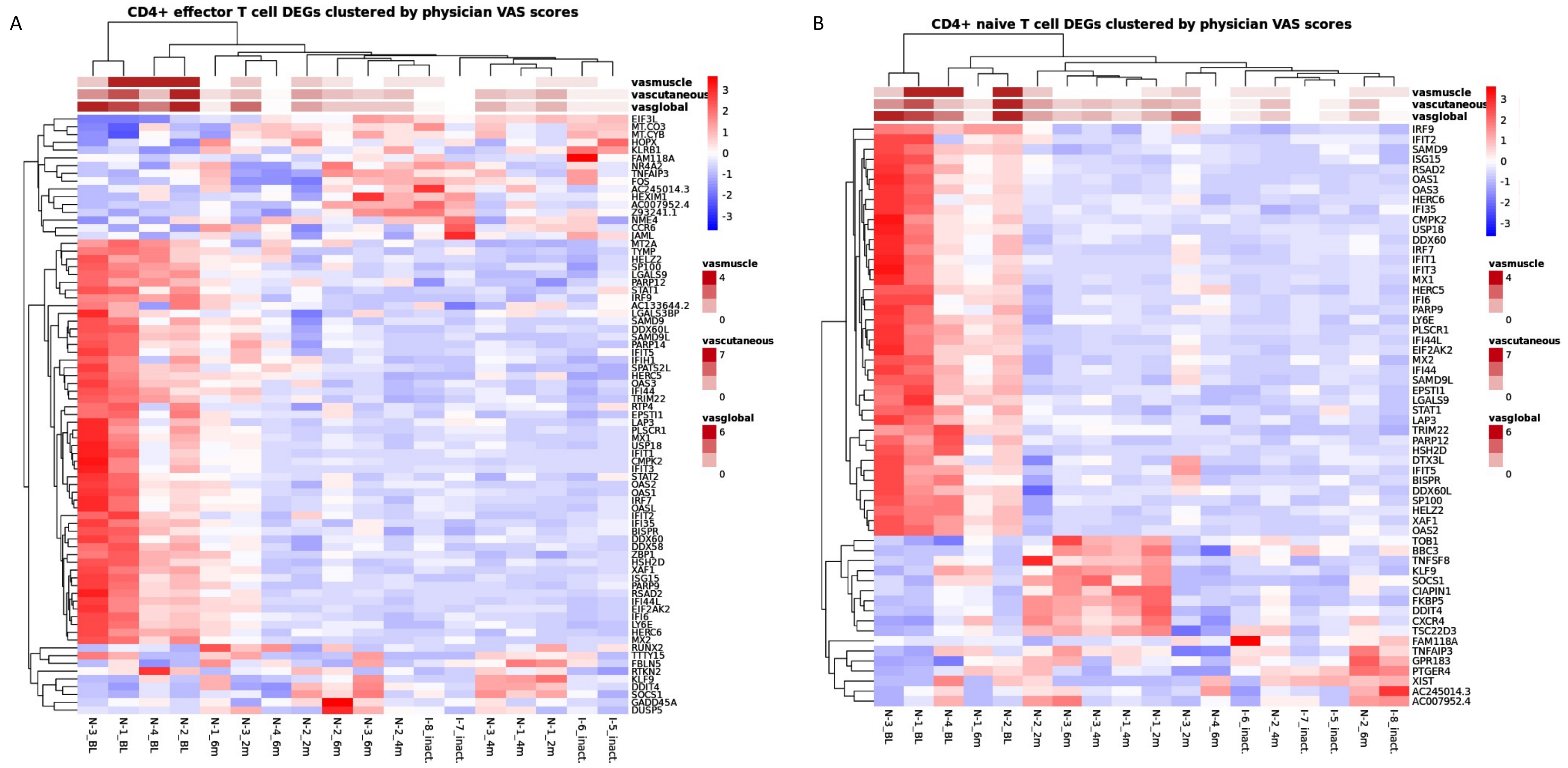


**Supplementary Fig. 10:** Heatmaps displaying hierarchical clustering of the psuedobulk expression of the union of genes differentially expressed in both longitudinal and cross-sectional analyses in CD4+ effector T cells (A) and CD4+ naïve T cells (B). Samples are annotated by disease activity measures including physical VAS scores for global ("vasglobal"), cutaneous ("vascutaneous"), and muscle ("vasmuscle") domains.

### Supplementary Figure 11

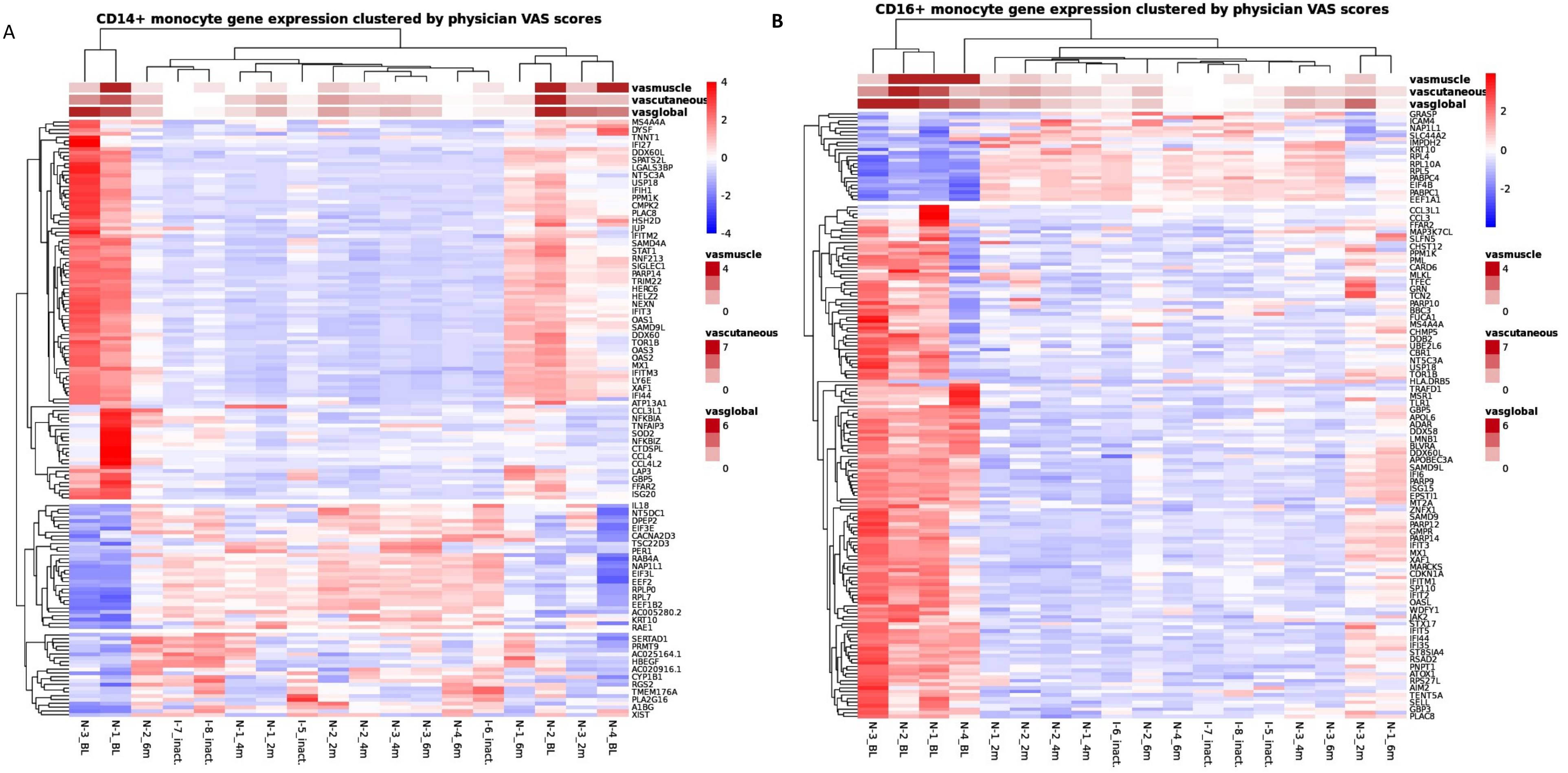


**Supplementary Fig. 11:** Heatmaps displaying hierarchical clustering of the psuedobulk expression of the intersection of genes differentially expressed in both longitudinal and cross-sectional analyses in CD14+ monocytes (A) and CD16+ monocytes (B). Only half of the genes are annotated for visualization purposes. Samples are annotated by disease activity measures including physical VAS scores for global (“vasglobal”), cutaneous (“vascutaneous”), and muscle (“vasmuscle”) domains.

### Supplementary Figure 12

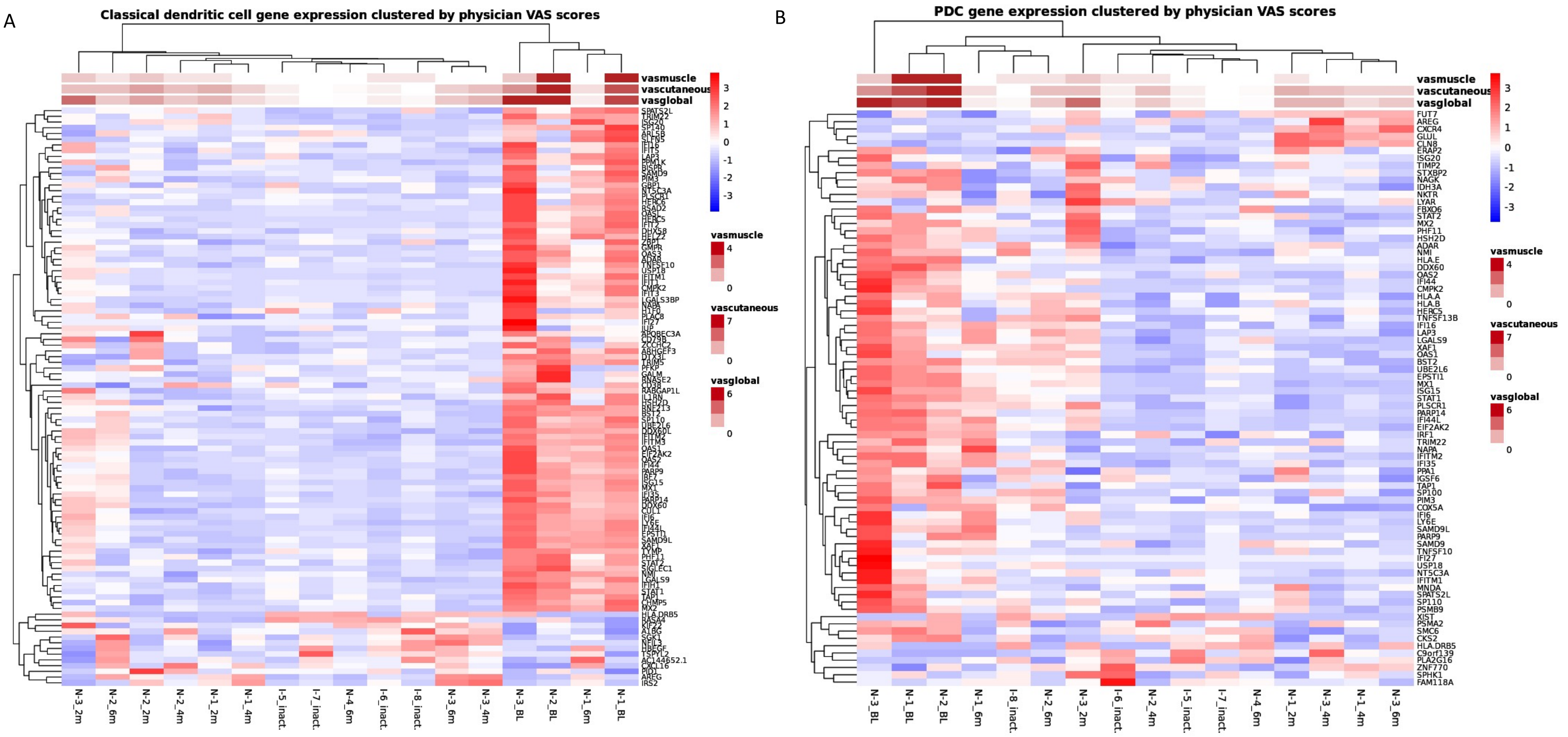


**Supplementary Fig. 12:** Heatmaps displaying hierarchical clustering of the psuedobulk expression of the genes differentially expressed in both longitudinal and cross-sectional analyses. The set of genes in classical dendritic cells is the intersection of genes expressed in both analyses (A) and the set of genes in plasmacytoid dendritic cells (PDC) is the union of genes expressed in both analyses (B). Samples are annotated by disease activity measures including physical VAS scores for global ("vasglobal"), cutaneous ("vascutaneous"), and muscle ("vasmuscle") domains.

### Supplementary Figure 13

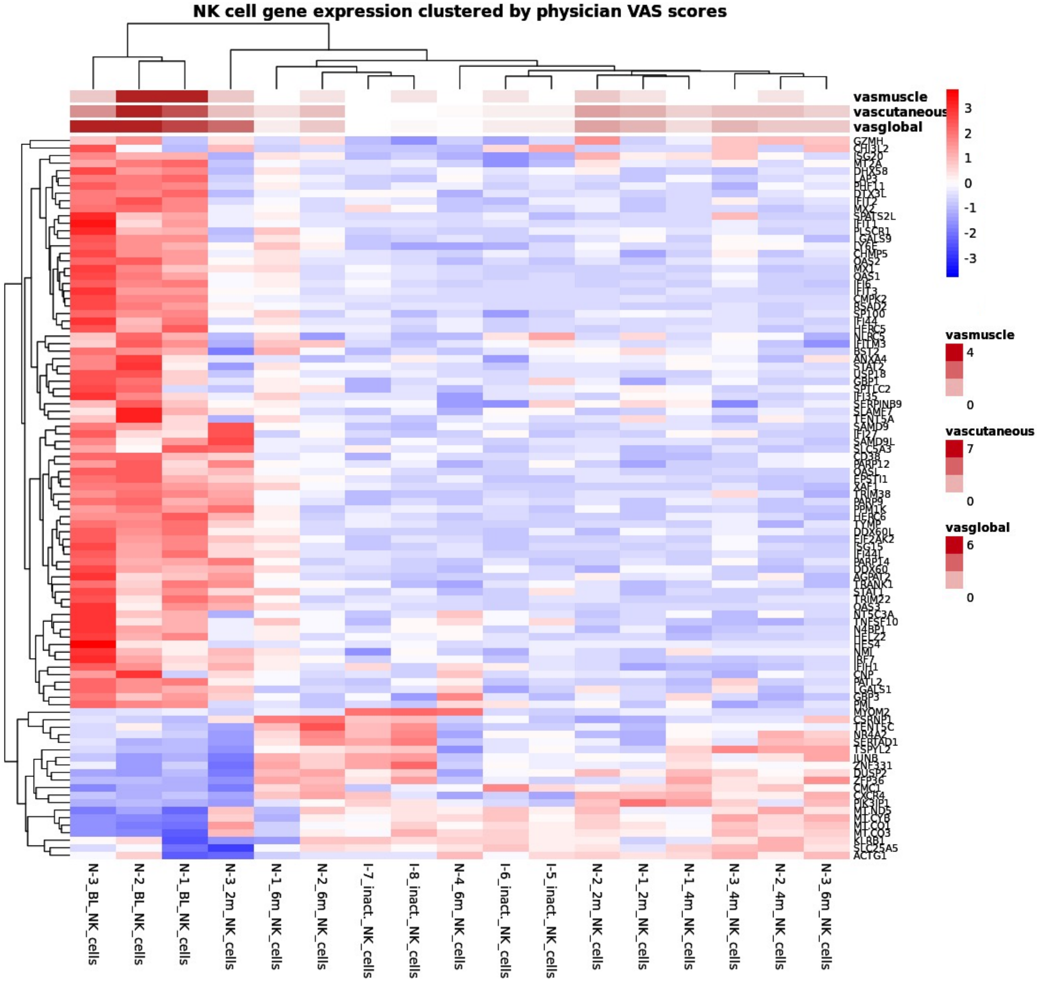


**Supplementary Fig. 13:** Heatmaps displaying hierarchical clustering of the psuedobulk expression of differentially expressed genes in all NK cells. The gene set is intersection of the union of genes differentially expressed longitudinally in both CD56dim NK and CD56bright NK cells and the union of genes differentially expressed cross-sectionally in both CD56dim NK and CD56bright NK cell populations. Samples are annotated by disease activity measures including physical VAS scores for global ("vasglobal"), cutaneous ("vascutaneous"), and muscle ("vasmuscle") domains.

#### Supplementary Figure 14: Module Reactome Pathway Enrichment

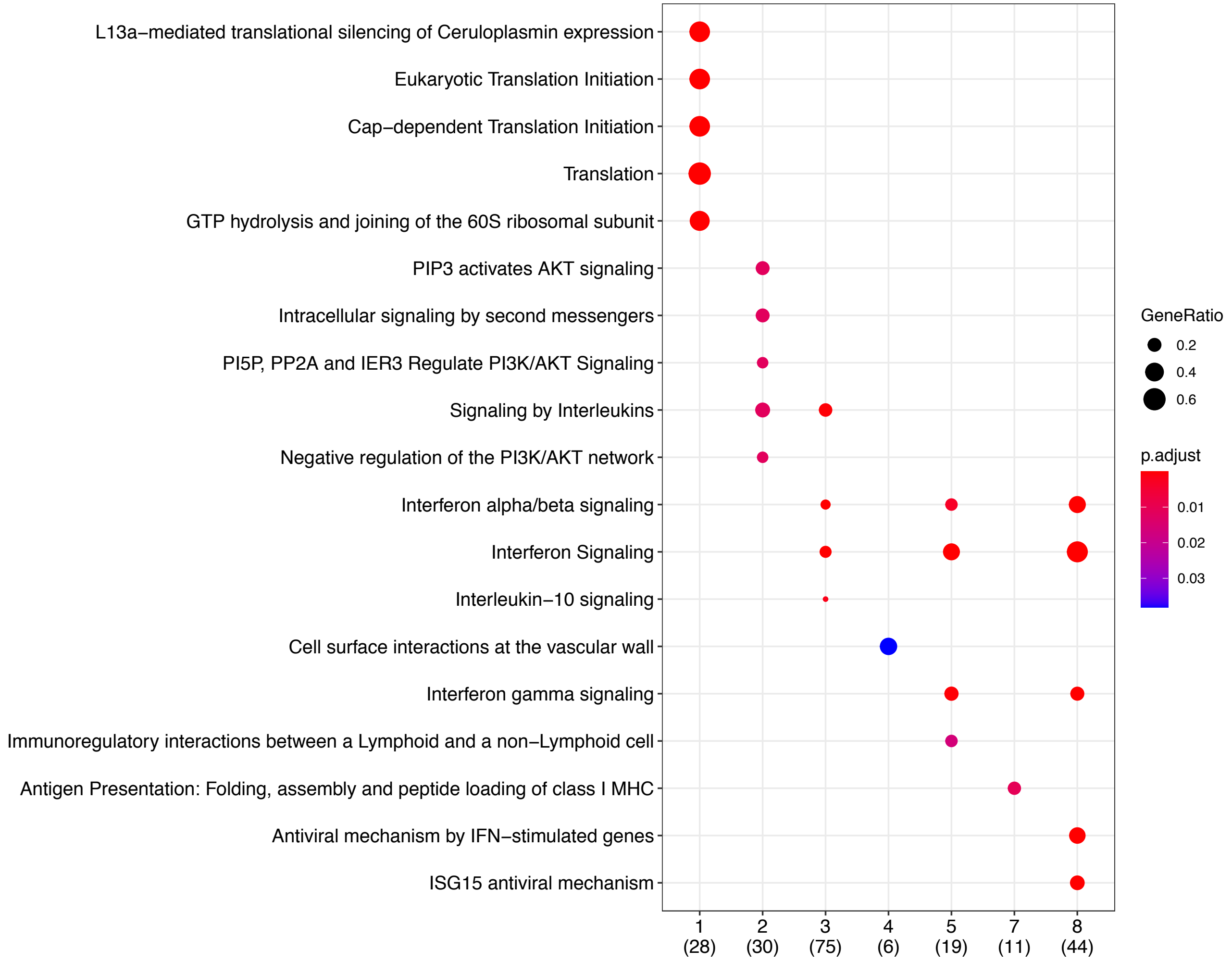


**Supplementary Fig. 14:** Enrichment of Reactome pathway terms by module using ClusterProfiler. There were no enrichment terms for modules 6, 9 and 10.

### Supplementary Figure 15

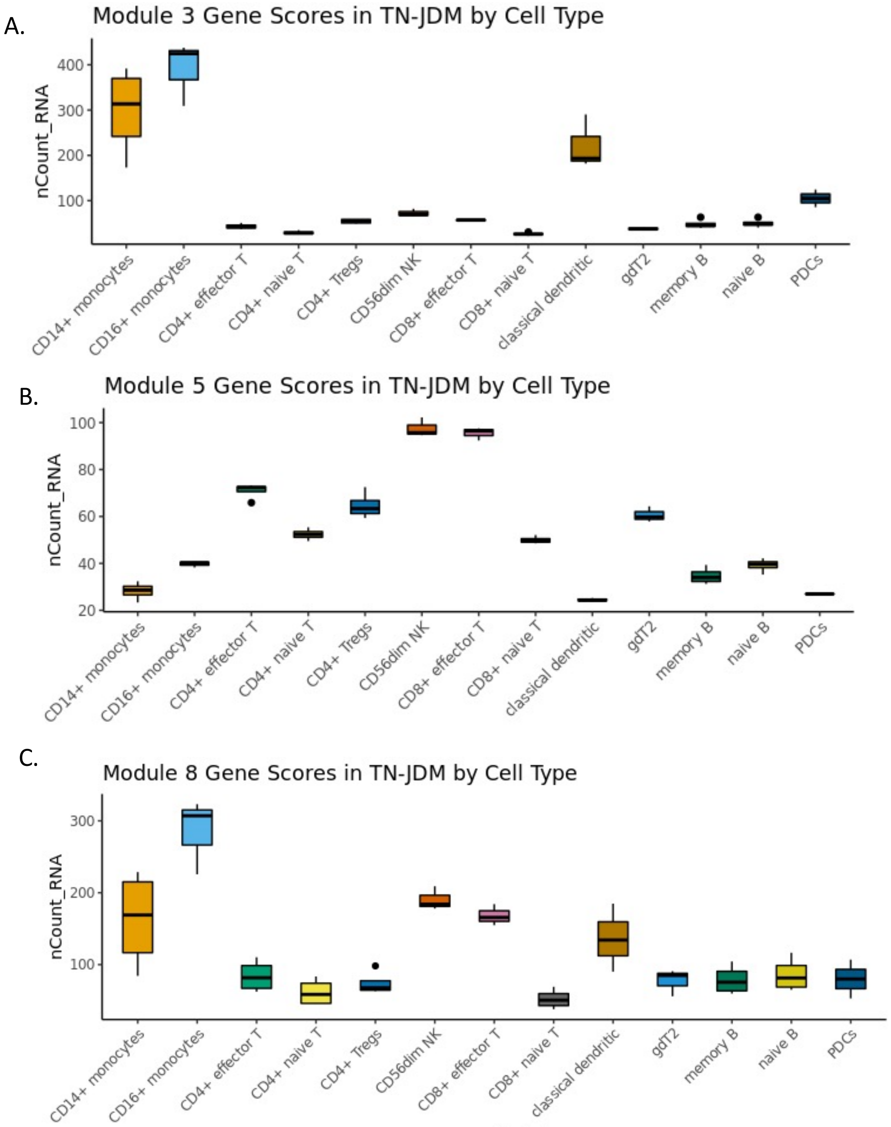


**Supplementary Fig. 15:** Average gene scores for each cell type in treatment-naïve patients for modules 3 (A), 5 (B), and 8 (C).
